# Genetic variants associated with 90-day survival in sepsis patients

**DOI:** 10.64898/2026.11.23.26368566

**Authors:** Zeggane Syphax, Mslmane Alaa, Fleuriet Jérôme, Heming Nicholas, Bacq-Daian Delphine, Olaso Robert, Deleuze Jean-François, Annane Djillali, Garchon Henri-Jean, RHU RECORDS

## Abstract

Sepsis is a highly prevalent and life-threatening syndrome resulting from an exaggerated inflammatory response to an infection. Its etiology is multifactorial and involves both environmental and genetic factors that are presently poorly understood. We conducted a genome-wide association study (GWAS) on sepsis patients’ survival at 90-day following admission in the ICU. We extracted genotypes of single-nucleotide variants (SNVs) from whole-genome sequencing data of 1030 patients from three cohorts, including APROCCHSS, RECORDS-Observational and TRIAL. We used a proportional hazard Cox model adjusted for age, the first two genetic principal components and the cohort. We identified 7 significant SNVs that reached genome-wide significance (*P* < 5 × 10⁻⁸). One of them, rs1474684, maps to the last intron of the phospholipase C Beta 1 (*PLCB1*) gene and likely is causative, following fine-mapping. Two other SNVs map to *IMPDH1P7* gene for rs146038228 and *CALCRL*-*AS1* for rs4667156. Both genes lie in a 400 kb region showing extended linkage disequilibrium that prevented resolution of the causative variant. The other four variants were low frequency SNVs mapping to non-coding or intergenic regions near *ENSG00000288587* (rs11754929), *ENSG00000248172* (rs74570843), *ISG20* (rs139793868), and *RNU6-144P* (rs78812419). Gene Set Enrichment Analysis, a sensitive nonparametric method to rank pathways associated with variants and genes, consistently highlighted terms associated with the central nervous system, synapse physiology and cardio-endocrine regulation. Altogether, our findings shed new light on genetic factors and pathways associated with survival in sepsis patients. However, further investigation in an independent cohort will be necessary for validation of the associations.

## Introduction

Sepsis is a life-threatening multifactorial disease resulting from a dysregulated inflammatory response to infection^1^. It affects 166 million people worldwide every year, and is responsible for 21.4 million deaths^2^. Moreover, the quality of life of the survivors is significantly impaired, with half suffering from long-term sequelae, particularly neurological and cognitive deficits^3^. In 2017, the World Health Organization (WHO) declared the improvement of sepsis prevention, recognition, and treatment a global health priority and the most urgent unmet medical need of our times, reaffirming this call to action in 2020^4^.

Genetic variation has been demonstrated to play a key role in shaping the host’s immune response to infection^5^. In the past two decades, genome-wide association studies (GWAS) have provided a robust framework to uncover the common genetic factors involved in predisposition to disease with complex inheritance^6,7^. Nonetheless, very few such studies have been conducted in sepsis specifically^8–15^. Moreover, these studies did not reveal any recurrent associated locus.

In this study, we aimed to characterize the genetic basis of sepsis outcomes. We conducted a GWAS on patient survival at ninety days in 1030 sepsis patients from three cohorts, including APROCCHSS^16,17^ (n=254), a double-blind randomized clinical trial, RECORDS-Observational^18^ (n=323), an observational COVID-19 cohort, and TRIAL^19^ (n=453), an ongoing adaptive double-blind randomized trial. Genotypes were obtained through whole-genome sequencing, thus avoiding the pitfalls of imputation. Through this approach, we identified 7 single nucleotide variants (SNVs) significantly associated with genome-wide significance. These findings reveal novel sepsis pathogenesis pathways, offering new genomic insights for risk stratification, prognosis, and future therapeutic targeting.

## Results

### Late-survival analysis identifies seven genome-wide significant SNVs

We conducted a genome-wide survival analysis for 90-day mortality comprising 1030 patients (743 survivors, 287 non-survivors) from 3 distinct cohorts (Table 1, Supplementary Figure 1a,b). Genomic inflation was adequately controlled (λ = 1.009). We identified seven single nucleotide variants (SNVs) across six distinct genomic loci that reached genome-wide significance (*P* < 5 × 10⁻⁸, Fig. 1a, Table 2). Forest plots of effect sizes (Hazard Ratio, HR) of each SNV by cohort confirmed the direction of effects and highlighted the consistency of the results (Fig. 1b).

**Figure 1.**
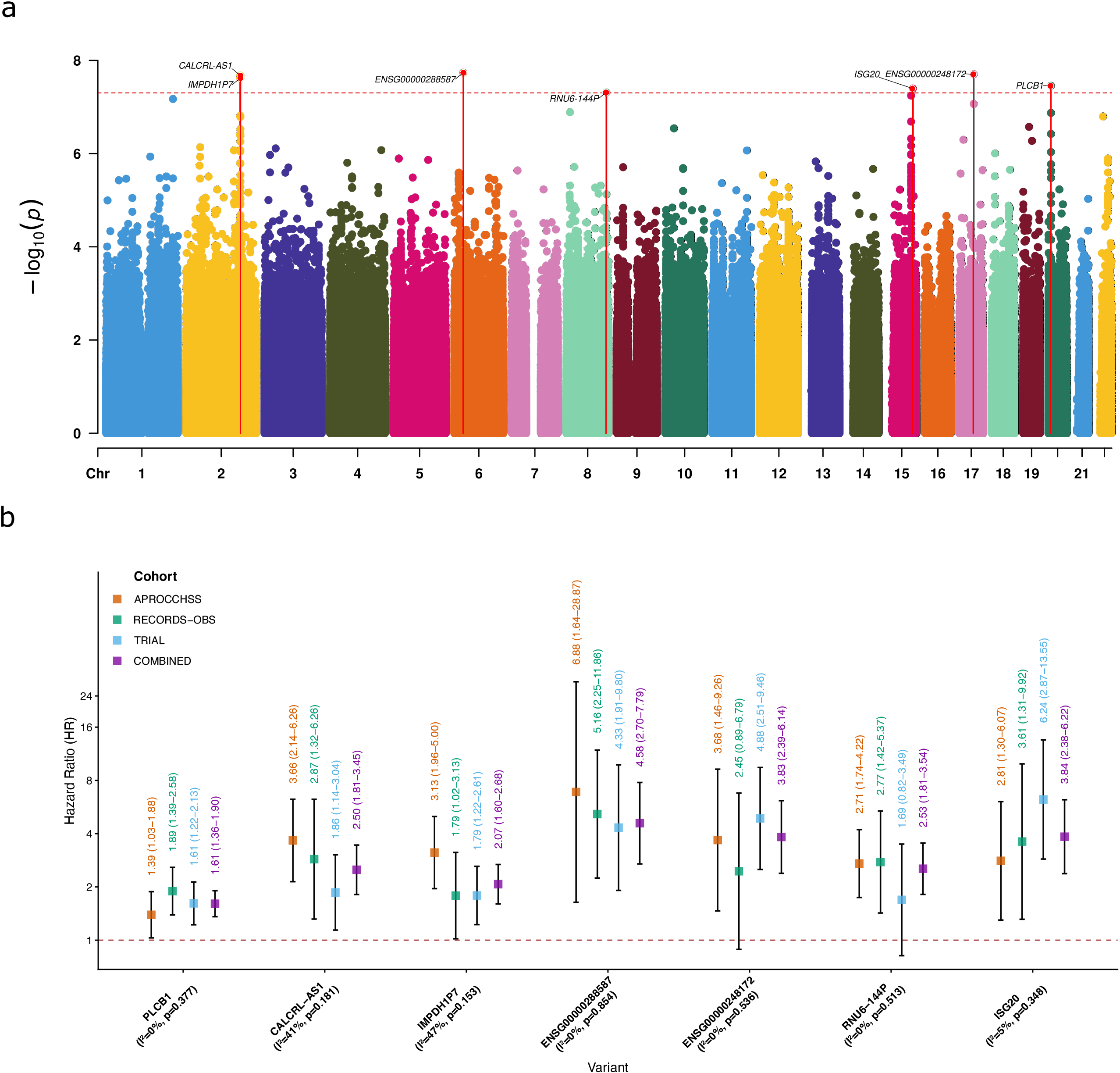
Genome-wide associations with variant effect sizes by cohort for 90-day sepsis survival. (a) Manhattan plot of genome-wide associations with 90-day survival in sepsis patients. The statistical significance −log₁₀(P) (y-axis) is plotted against the genomic position of variants (chromosomes 1 to 22, on the x-axis). The horizontal dashed red line indicates the genome-wide significance threshold (P = 5 × 10⁻⁸). Red dots above this threshold represent significantly associated variants, connected by solid red vertical lines to their respective chromosomal positions and annotated with the closest gene. (b) Forest plot of hazard ratios by cohort for associated variants. Colored squares represent the HR (y-axis) and 95% confidence intervals (CI) for individual cohorts (APROCCHSS, RECORDS-OBS, and TRIAL) and the main analysis. Horizontal dashed red line indicates the null effect (HR = 1). Solid vertical lines represent 95% CI. X-axis shows variants by their closest gene, alongside heterogeneity score (*I*^2^) and Cochrane’s Q statistic (p-value).

**Table 1:**
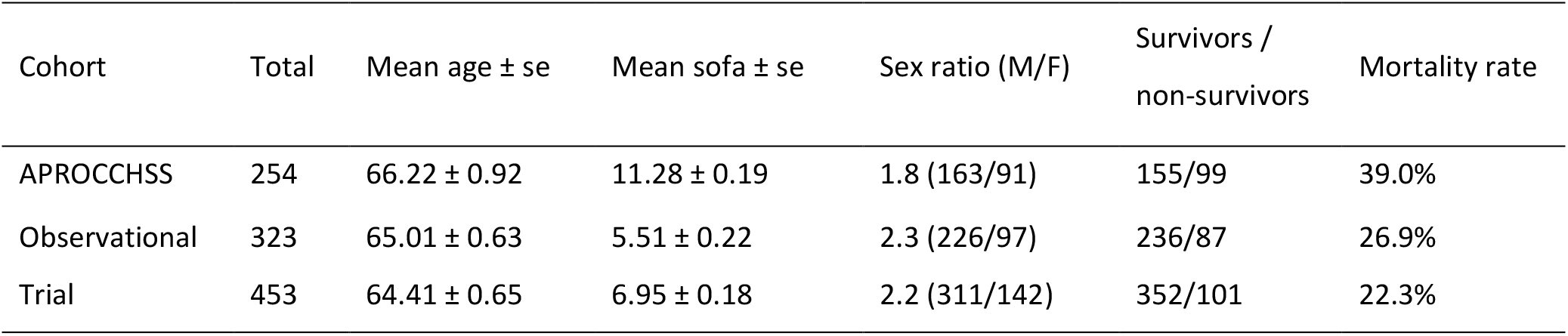
Main characteristics of the three study cohorts.

**Table 2:** Genome-wide significant variants associated with 90-day sepsis survival.

| RsID | Position (chr:distance) | Alt/Ref | MAF | Gene | Gene Func. | HR (95% CI) | *P* value |
|---|---|---|---|---|---|---|---|
| rs146038228 | 2:186951556 | T/C | 0.078 | IMPDH1P7 | Pseudogene | 2.07 (1.60-2.68) | 2.4 × 10<sup>−8</sup> |
| rs4667156 | 2:187232732 | G/A | 0.039 | CALCRL-AS1 | lncRNA | 2.50 (1.81-3.45) | 2.1 × 10<sup>−8</sup> |
| rs11754929 | 6:31460992 | T/C | 0.014 | ENSG00000288587 | lncRNA | 4.58 (2.70-7.79) | 1.8 × 10<sup>−8</sup> |
| rs78812419 | 8:137135567 | C/T | 0.028 | RNU6-144P | snRNA | 2.53 (1.81-3.54) | 4.9 × 10<sup>−8</sup> |
| rs139793868 | 15:88678201 | G/C | 0.012 | ISG20 | Protein coding | 3.84 (2.38-6.22) | 4.0 × 10<sup>−8</sup> |
| rs74570843 | 17:50021979 | T/C | 0.014 | ENSG00000248172 | lncRNA | 3.83 (2.39-6.14) | 2.0 × 10<sup>−8</sup> |
| rs1474684 | 20:8795913 | G/A | 0.323 | PLCB1 | Protein coding | 1.61 (1.36-1.90) | 3.5 × 10<sup>−8</sup> |

Two association signals were characterized by well-defined linkage disequilibrium (LD) structure. On chromosome 2, the lead variant rs4667156 (MAF= 0.04, *P* = 2.1 × 10⁻⁸), located in the lncRNA *CALCRL-AS1* gene and antisense to both *CALCRL* and *TFPI*, was significantly associated with increased mortality for carriers of the minor allele (HR = 2.50, 95% CI: 1.81-3.45, Supp. Fig. 2a). Regional association plots revealed an extended LD block encompassing some 400 Kb and including 5 genes and 75 suggestively associated variants, alongside a secondary genome-wide significant signal mapping to *IMPDH1P7* (rs146038228, MAF = 0.08, *P* = 2.4 × 10⁻⁸) in weak LD with the index SNV (r² = 0.436, Fig. 2a, Supp. Fig. 2b). On chromosome 20, we detected a prominent association peak mapping to the last intron of the *PLCB1* gene, driven by the common variant rs1474684 (MAF = 0.32, *P* = 3.5 × 10⁻⁸, Fig. 2b). Survival trajectories confirmed a dose-dependent reduction in survival for risk-allele carriers at this locus (HR = 1.61, 95% CI: 1.36-1.90, Supp. Fig. 2c).

**Figure 2.**
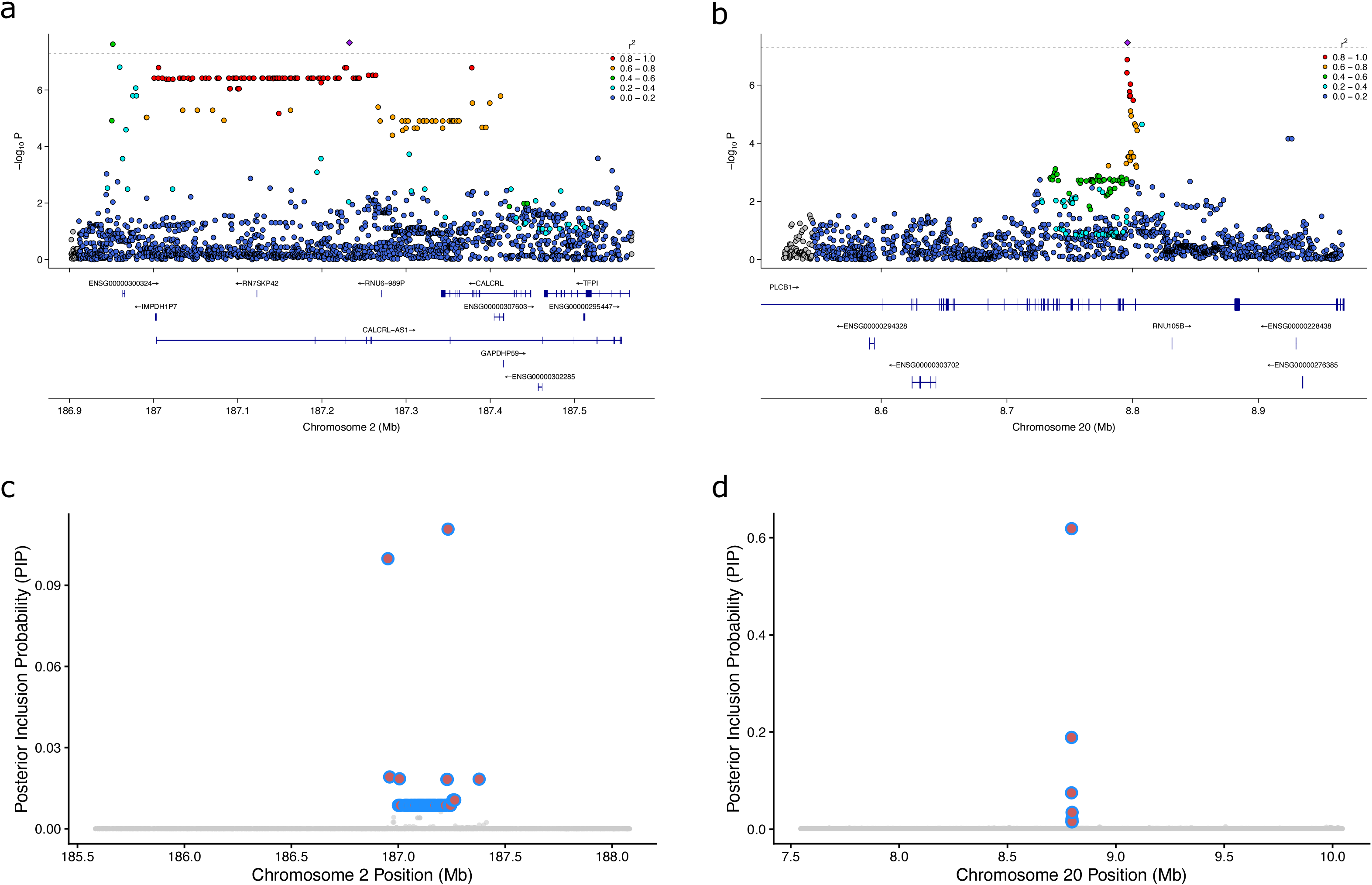
Regional association plots and fine-mapping of *CALCRL-AS1* and *PLCB1* loci. (a) Locus zoom plot detailing the associated variants at *CALCRL-AS1* genomic region. The top panel displays the association values −log₁₀(P) (y-axis) of SNVs according to their chromosomal position (x-axis). The top index variant at each locus is highlighted as a purple diamond. Surrounding variants are color-coded to reflect their linkage disequilibrium (r²) with the index variant (red, 0.8–1.0; orange, 0.6–0.8; green, 0.4–0.6; light blue, 0.2–0.4; dark blue, 0.0–0.2). The horizontal dashed grey line denotes the genome-wide significance threshold (P = 5 × 10⁻⁸). The bottom panels illustrate the local gene annotations and exon-intron structures, with arrows indicating the direction of transcription. (b) Locus zoom plot detailing the associated variants at *PLCB1* genomic region. (c) Credible set (CS) resulting from the fine-mapping of *CALCRL-AS1* locus. The y-axis shows the PIP (Posterior Inclusion Probability). The x-axis displays the variants according to their position. Variants that belong to the CS are circled in blue. CS includes seventy-five variants, with an average LD within the CS of (*r*^2^ = 0.956). (d) Credible set resulting from the fine-mapping of *PLCB1* locus. CS includes seven variants, with an average LD of (*r*^2^ = 0.896).

The other four identified regions showed isolated association signals, driven by low-frequency variants (MAF < 0.03), and characterized by sparse local linkage disequilibrium (LD). These included regions on chromosome 6 near the novel transcript *ENSG00000288587* (rs11754929, *P* = 1.8 × 10⁻⁸, HR = 4.58), on chromosome 17 near the lncRNA *ENSG00000248172* and *DLX3* (rs74570843, *P* = 2.0 × 10⁻⁸, HR = 3.83), chromosome 15 near *ISG20* (rs139793868, *P* = 4.0 × 10⁻⁸, HR = 3.84), and chromosome 8 near the *RNU6-144P* pseudogene and *FAM135B* (rs78812419, *P* = 3.5 × 10⁻⁸, HR = 2.53). Kaplan-Meier trajectories for these four variants demonstrated significantly reduced 90-day survival for carriers of the risk alleles (Supp. Fig. 2d-g). Regional association plots confirmed the absence of extended LD blocks, and conditional association testing adjusting for the respective index SNVs (lead SNVs) revealed no independent secondary signals at these loci (Supp. Fig. 3a-d).

Because the resolution of causal variants requires robust local LD support, we prioritized the topologically robust loci at chromosome 2 (*CALCRL-AS1*) and chromosome 20 (*PLCB1*) for subsequent statistical fine-mapping and molecular colocalization analyses.

### Statistical fine-mapping and expression colocalization

We first used the Sum of Single Effects (SuSiE) model. At the chromosome 2 locus (*CALCRL-AS1*), the credible set (CS) remained large, including 75 SNVs (Fig. 2c). A very dense and unfragmented LD block surrounding the index SNV (Supp. Fig. 4a) prevented the algorithm from confidently isolating a limited set of causal variants (maximum Posterior Inclusion Probability [PIP] < 0.12, Supp. Table 1). In contrast, at the chromosome 20 locus (*PLCB1*), we identified a CS of only 7 variants (Fig. 2d). Despite strong average LD (mean *r*^2^ = 0.89), the regional correlation matrix revealed a fragmented structure that allowed the model to successfully prioritize the index SNV as potentially the primary causal candidate (Supp. Fig. 4b). This SNV showed a PIP of 61.8%, while the secondary variant achieved a PIP of only 18.8% (Supp. Table 2).

To determine whether these associations are driven by genetic regulation of local gene expression and splicing, we performed cis-eQTL (expression quantitative trait locus) and cis-sQTL (splicing quantitative trait locus) mapping followed by colocalization analyses. We analyzed RNA sequencing data from whole blood for a subgroup of 279 patients (227 survivors vs 52 non-survivors) to uncover sQTLs and eQTLs at ±500 Kb flanking each index variant of both loci (Supp. Tables 3-6). We then assessed the colocalization of the GWAS late-survival signal with the expression and splicing profiles of both loci and their neighboring genes. However, neither at the chromosome 2 nor at the chromosome 20 locus, there was evidence for a shared causal variant (H4) or distinct causal variants (H3) for any transcript in the region (Supp. Tables 7-10). Together, these findings reveal that the genetic variants driving sepsis survival at these loci do not heavily influence baseline transcript abundance or structural splicing in circulating leukocytes. The causal mechanisms might thus operate through other QTLs, such as chromatin accessibility, in distinct tissues or require specific physiological stimulation.

### Gene prioritization and pathway analysis identify systemic molecular mechanisms

To contextualize the broader genetic information of the GWAS, we transitioned from single variant mapping to a genome-wide functional enrichment approach. We performed Gene Set Enrichment Analysis (GSEA) by prioritizing genes based on the statistical significance of their top associated variant (ranked by minimum P-value). Within the resulting cnetplots, we added the −*log*_10_(*HR*) of each gene as a color indicator to spot potential clusters of pathways that could be driven predominantly by protective (blue, HR < 1) or risk-increasing (red, HR > 1) genetic effects. Notably, this visualization revealed no distinct protective or risk-increasing pathways (Fig. 3). Interrogating the Gene Ontology (GO), Kyoto Encyclopedia of Genes and Genomes (KEGG), and Reactome databases revealed a highly convergent overall genetic architecture (Fig. 3, Supp. Fig. 5). Strikingly, the most prominently enriched terms were driven by neuronal and synaptic biology. GO analysis was heavily dominated by nervous system development and trans-synaptic signaling, including terms such as axon development, synapse organization, and regulation of neuron projection development (Fig. 3a). This neuro-centric signature was fully recapitulated in the KEGG and Reactome networks, which identified massive enrichments of terms for the neuronal system, transmission across chemical synapses, neuroactive ligand-receptor interaction, and specific neurotransmitter axes, notably glutamatergic and GABAergic synapses (Fig. 3b-c).

**Figure 3.**
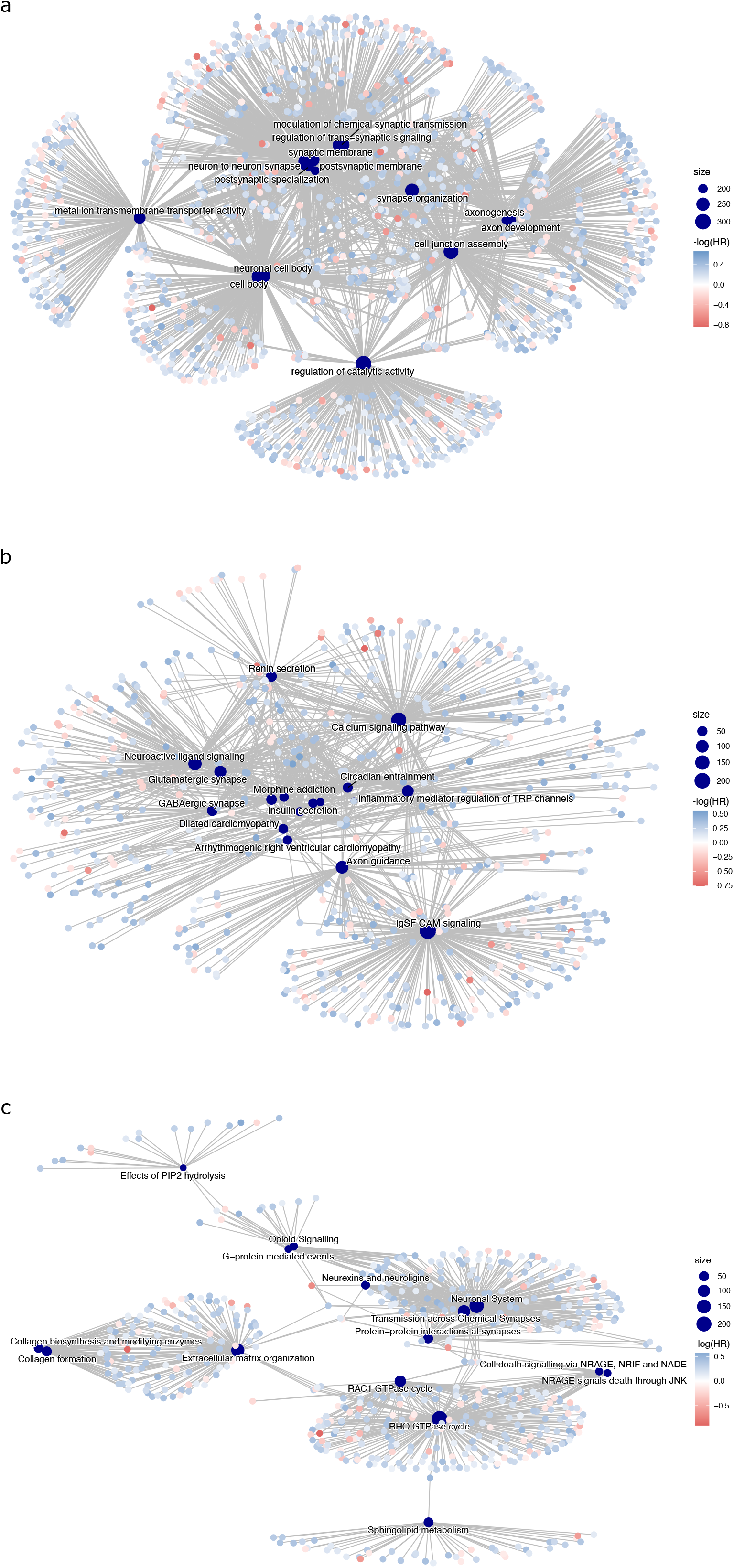
Gene-concept network plots (cnetplots) of enriched biological pathways associated with sepsis survival. Gene Set Enrichment Analysis (GSEA) networks relating pathways with genes across three annotation databases: Gene Ontology/GO (a), KEGG (b), and Reactome (c). Central dark blue nodes represent significantly enriched pathways or functional terms, with the size of the node proportional to the number of genes included in the specific category. Smaller peripheral nodes represent the individual genes driving these pathway enrichments. Grey edges connect common genes to multiple pathways, illustrating functional overlap and pleiotropy. Gene nodes are color-coded based on their association statistic representing the − log(HR), where blue (positive values, HR < 1) indicates a protective effect on survival, and red (negative values, HR > 1) indicates an increased risk of mortality.

Beyond neuronal and synaptic biology, the pathway analysis highlighted involvement of cardiovascular, endocrine system, and cell structure integrity. KEGG pathways revealed significant enrichments for systemic hemodynamic regulators, including renin secretion, aldosterone synthesis and secretion, and calcium signaling pathways, alongside structural cardiac conditions such as dilated cardiomyopathy (Fig. 3b). In parallel, Reactome analysis underscored the importance of vascular and endothelial stability, isolating pathways involved in extracellular matrix organization, collagen biosynthesis, and sphingolipid metabolism. We also observed significant enrichment for RAC1 and CDC42 GTPase cycles (Fig. 3c), which are critical regulators of cytoskeletal dynamics and endothelial barrier function during severe inflammation.

Collectively, these networks indicate that the genetic variance governing 90-day survival in sepsis extends far beyond classical immune cell function, relying heavily on host neuronal-inflammatory crosstalk, vascular integrity, and neuro-endocrine resilience.

## Discussion

We identified 7 SNVs associated with survival of sepsis patients at 90-day with genome-wide statistical significance and mapping to 6 distinct chromosomal regions. The fact that the associations showed no evidence of heterogeneity across the three cohorts of the study, of which two are part of a double-blind randomized controlled clinical trial, indicates that the findings are robust, although we acknowledge the necessity of a validation in an independent cohort. Moreover, the genotypes were directly observed from whole-genome sequencing data rather than imputed. Indeed, imputation is almost always required as genotypes are most often obtained using microarrays but remains limited by the reference population considered. By avoiding imputation, our use of WGS data therefore also increases the confidence in our findings.

As is common in multifactorial diseases, all the SNVs we detected mapped to noncoding regions. At chromosome 20, the lead SNV rs1474684 mapped to the 11^th^ and last intron of the *PLCB1* gene and fine-mapping revealed that it very likely is the causative variant. *PLCB1* encodes phospholipase C beta 1, a critical calcium-dependent enzyme that hydrolyzes membrane phospholipids to drive intracellular signal transduction. Crucially, the PLCB1 protein acts as a core downstream effector for G protein-coupled receptors (GPCRs) and glutamatergic signaling cascades. In clinical genetics, pathogenic mutations in *PLCB1* are robustly linked to a spectrum of severe, early-onset epileptic encephalopathies. This is consistent with our term-enrichment analysis revealing pathways such as synaptic organization, trans-synaptic chemical signaling, and calcium flux pathways. This suggests that PLCB1-dependent signaling networks driving neuronal excitability might be involved in sepsis neurological dysfunctions such as sepsis-associated encephalopathy (SAE), which frequently complicates systemic infections. The molecular mechanism of action of rs1474684 remains to be elucidated. It did not alter the expression level or the splicing of transcripts of *PLCB1* and neighboring genes. However, the QTLs might be tissue-specific and expressed elsewhere, including in the CNS or the cardiovascular system. Other mechanisms are also possible.

On chromosome 2, the extended LD at *CALCRL-AS1* and neighboring loci prevented the statistical resolution to a single causal variant. The long non-coding RNA *CALCRL-AS1* is transcribed antisense to two critical regulators of vascular homeostasis: *CALCRL* and *TFPI*. The *CALCRL* gene encodes the calcitonin receptor-like receptor, the primary signaling receptor for adrenomedullin (ADM), a potent vasodilator and endothelial-protective peptide currently under extensive clinical investigation as a therapeutic target in sepsis. Moreover, serum level of ADM was recently associated with 90-day survival^20^. However, we did not identify a significant effect of *CALCRL-AS1* index variant (rs4667156) on plasma ADM concentrations in a subset of the TRIAL cohort (Supp. Fig. 6). In parallel, tissue factor pathway inhibitor (*TFPI*) is a master regulator of the coagulation cascade, protecting against the microvascular thrombosis and severe endothelial damage that characterize systemic hyperinflammation^21^. Notably, pathogenic variants within the *CALCRL-AS1* locus have been previously implicated in lymphatic malformation 8.

Gene set enrichment analyses provide a sensitive approach to detect weak signals by pooling genes belonging to the same pathway and ranking them in the whole dataset. In our case, this assumes that the expression of the genes is effectively altered by the variants mapping to them, which is difficult to verify. It remains that our analysis consistently highlighted terms implicating the central nervous system. This is in line with the neurological disorders caused by pathogenic variants in the *PLCB1* gene.

Our failure to replicate previously published variants (Supp. Table 11) can be explained by several factors, notably including differences in the study design (case-control), phenotype definition, or in the endpoint (28-day), or a lack of power for rare variants. This warrants studying additional independent cohorts, hopefully in an international setting.

## Materials and methods

### Study population

Detailed information regarding the design, protocols, and statistical analysis plans of the APROCCHSS, RECORDS-OBS, and TRIAL cohorts has been previously documented^16,18,19^. The trial protocols were approved by the Ethics Committee (CPP) of Dijon, France on April 6, 2020 and The Agence Nationale pour la Sécurité du Médicament on April 9,2020. Written informed consent was obtained from all participants or their legally authorized representatives, and where applicable, deferred written informed consent was obtained. All authors were granted complete and independent access to the data, and they affirm the integrity, accuracy, and comprehensiveness of the data and analyses, as well as adherence to the study protocol.

This study utilized WGS data from 1065 patients enrolled across these three cohorts as part of the hospital-university research projects RHU RECORDS and the IHU SEPSIS. Prior to sequencing, the genetic sex of all samples was verified via PCR. Samples exhibiting discordance between PCR-determined sex and clinical metadata were excluded and consequently not sequenced. After exclusion of 22 patients due to consent withdrawal or missing metadata, and 13 patients due to relatedness or duplicated samples identified using default parameters of KING^25^ (kinship >0.17), the final genomic analysis comprised 1030 patients. The final study included 254 patients from the APROCCHSS double-blind randomized clinical trial, 323 patients from the RECORDS-OBS observational COVID-19 cohort, and 453 patients from the ongoing RECORDS TRIAL adaptive double-blind randomized trial (Table 1). The clinical phenotype for the genome-wide survival analysis was defined as time-to-death at 90-day following inclusion.

### Sequencing, workflow and work environment

Total DNA was isolated from the whole blood collected in EDTA tubes, utilizing the Monarch® Spin gDNA Extraction Kit (New England Biolabs) according to the manufacturer’s protocols. DNA concentration was measured with a LabChip® DS. Samples were qualified and libraries were prepared with Illumina TruPath. They were sequenced with an Illumina NovaSeq 6000 sequencer at the National Center for Research in Human Genomics (CNRGH). The distribution of mean sequencing depth and genome coverage was adequate in all samples (Supp. Tables 12-13, Supp. Fig. 7).

Studies were operated in a DELL R940 server with OS DEBIAN 12.15, and R version 4.5.3.

Raw sequencing reads were aligned to the human reference genome (GRCh38) using *bwa-mem*, and variant calling was performed using *Haplotype caller*^22^ following the Genome Analysis Toolkit (GATK v4.6.1.0) Best Practices workflow to generate genomic VCFs (*gVCFs*). These *gVCFs* were subsequently subjected to rigorous quality control and joint genotyping. Prior to processing, genomic regions known to produce artifactual alignments were excluded using the ENCODE curated blacklist (*GRCh38_unified_blacklist.bed*, ENCFF356LFX). Left-alignment and multiallelic normalization of the variants were performed using *bcftools*^23^ (v1.19) *norm*. The normalized *gVCFs* were then aggregated into genomic databases partitioned by chromosomal region using GATK *GenomicsDBImport*. Joint variant calling was executed across the samples using GATK *GenotypeGVCFs* against the GRCh38 reference genome (hs38DH.fa), generating chromosome-level raw VCFs that were subsequently concatenated using GATK *MergeVcfs*.

To eliminate false-positive variant calls, a hard filter was first applied to remove sites with excessive heterozygosity (ExcessHet > 54.69). Next, we applied the GATK Variant Quality Score Recalibration (VQSR) pipeline. VQSR was trained and applied separately for SNPs and INDELs to calculate the Variant Quality Score Log-Odds (VQSLOD). For INDEL recalibration, the Gaussian mixture model (maximum of 4 Gaussians) was trained using Fisher Strand (FS), ReadPosRankSum, MQRankSum, Quality by Depth (QD), Strand Odds Ratio (SOR), and Depth (DP). Training sets for INDELs included the Mills and 1000G gold standard, Axiom Exome Plus, and dbSNP 146. For SNP recalibration (maximum of 6 Gaussians), the model incorporated QD, MQRankSum, ReadPosRankSum, FS, Mapping Quality (MQ), SOR, and DP, utilizing HapMap 3.3, 1000G Omni 2.5, 1000G Phase 1 high-confidence SNPs, and dbSNP 146 as training resources. A strict truth sensitivity filter threshold of 99.0% was maintained for both variant types, corresponding to a minimum VQSLOD of 2.2709 for SNPs and −0.6209 for INDELs. Only variants assigned a PASS status were retained.

Global variant metrics and summary statistics were calculated using GATK *CollectVariantCallingMetrics* and *VariantEval*. Functional annotation was conducted using *bcftools*. Variants were annotated with identifiers from *dbSNP* (v156) and pathogenicity metrics from *CADD* (v1.7) and *ClinVar* (release 20231028). Allele frequencies were integrated from *gnomAD* (v4.1), and molecular consequences were predicted utilizing the Ensembl GRCh38.111 annotation with *bcftools CSQ*.

Finally, single-nucleotide variants (SNVs) were filtered using *PLINK*^24^ (v1.9) based on minor allele frequency (MAF > 1%), genotype missingness (< 1%), and Hardy-Weinberg equilibrium deviation (P > 0.0001), yielding a final set of 7,950,563 suitable SNVs.

### Principal Component Analysis and Population stratification

Reference population alleles were obtained from the 1000 Genomes Project (n = 3,202) and the Human Genome Diversity Project (HGDP; n = 948) and were intersected with the alleles from our dataset. PCA was subsequently conducted using *PLINK*, and the sepsis patient samples were projected onto the principal component space alongside the reference populations to visualize population stratification (Supp. Fig. 1a). To classify the genetic ancestry of the cohort, a k-nearest neighbors (KNN) algorithm was applied. The model was trained using the first ten principal components on the reference population alleles obtained from 1000GP and HGDP, and then was applied and tested on the first ten principal components of the alleles from our dataset. This assignment confirmed that the cohort was predominantly composed of Non-Finnish European (NFE) individuals, representing 82.43% of the samples (n = 849), followed by Middle Eastern (9.90%, n = 102) and African (4.76%, n = 49) ancestries (EAS = 11, AMR = 5, SAS = 5, Others = 8, Supp. Fig. 1c). To adjust for underlying genetic ancestry in the survival models, the first two principal components (PC1 and PC2) were used as covariates. The effect of genome-wide significantly associated SNVs was evaluated on each population (Supp. Figure 8, Supp Table 14).

### Genome-Wide Survival Analysis

GWAS of 90-day survival on common SNVs (MAF > 1%) was conducted on the aggregated cohorts (N = 1,030, comprising APROCCHSS n = 254, RECORDS-OBS n = 323, and TRIAL n = 453) with a Cox proportional hazard regression model using the R package *gwasurvivr*^26^, adjusting for age, cohort, and the first two principal components. For loci exceeding the genome-wide significance threshold (*P* < 5 × 10⁻⁸), local linkage disequilibrium (LD) architecture was calculated using *PLINK*. Regional association plots showing local LD structures were visualized using the R package *locuszoomr*^27^ with reference genes from the *Ensembl* database (v113).

### Statistical fine-mapping

We conducted statistical fine-mapping utilizing the Sum of Single Effects (SuSiE) model implemented via the R package *susieR*^28^ with default parameters. SuSiE is a Bayesian deconvolution method that uses LD and Z-scores to identify the minimum number of variants needed to explain the observed signal. For each locus of interest, linkage disequilibrium (LD) correlation matrices (±500 Kb flanking the index variant) were computed using *PLINK*. These locus-specific LD matrices were integrated with the corresponding GWAS summary statistics (Z-scores) to model potential causal signals (allowing for a maximum of L=10 causal variants). SuSiE then generated the 95% credible sets (CS) and calculated the posterior inclusion probability (PIP) for variants mapping within the associated loci.

### Transcriptomic Data Acquisition

Total RNA was isolated from whole blood collected in PAXgene tubes utilizing the Monarch Total RNA Miniprep Kit (New England Biolabs). RNA concentration and integrity were measured using a Quantus Fluorometer and a LabChip GX Touch nucleic acid analyzer (PerkinElmer), respectively. Samples with an RNA integrity number (RIN) > 7.0 were retained for sequencing. The final transcriptomic sub-cohort comprised 279 patients derived from the RECORDS-OBS (n = 24) and TRIAL (n = 255) cohorts. RNA sequencing was performed across three distinct genomic centers. For the subset sequenced at the Simone Veil Faculty genomic platform (UVSQ), libraries were prepared using the NEBNext rRNA Depletion Kit (New England Biolabs) and sequenced on an Illumina NextSeq 500/550, generating 75-bp paired-end reads. For subsets sequenced at the CNRGH and the Paris Brain Institute (ICM), libraries were constructed utilizing the Ribo-Zero Plus rRNA Depletion Kit (Illumina) and sequenced on an Illumina NovaSeq X Plus, yielding 100-bp paired-end reads.

Raw RNA-sequencing reads in FASTQ format were subjected to quality control utilizing *fastp* (v0.23.4) with default parameters to remove sequencing adapters and low-quality bases. The filtered reads were subsequently aligned to the human reference genome (GRCh38) utilizing the splice-aware aligner *STAR* (v2.7.11b). The alignment index was constructed leveraging the GENCODE v49 transcript annotation. To optimize data storage while preserving mapping integrity, the resulting alignment records were condensed into *CRAM* format. Gene-level expression quantification was executed concurrently with the alignment step utilizing *STAR’s* built-in quantification module. Read attribution was strictly resolved utilizing the reversely stranded counts (s2) to ensure accurate transcript assignment and prevent overlaps at polycistronic loci. Finally, the individual sample counts were parsed and aggregated into a unified raw gene expression matrix within the R statistical environment utilizing the *tidyverse* package suite.

### QTL mapping and colocalization Analysis

To identify quantitative trait loci (cis-eQTLs and cis-sQTLs) mapping to sepsis survival, regional WGS genotypes (±500 Kb flanking each genome-wide significant index variant) were extracted using *PLINK*. Matched whole-blood RNA-seq gene expression matrices were filtered to remove low-expressed transcripts (retaining genes with counts per million [CPM] > 0.01, corresponding to >1 read in >1% of patients). Filtered counts underwent Trimmed Mean of M-values (TMM) normalization, *log*2 transformation, and subsequent inverse normal transformation (INT) to enforce normally distributed residuals. To quantify alternative splicing events, intron excision ratios were calculated utilizing *LeafCutter*^29^ and identically subjected to INT. To account for hidden confounding variables and technical batch effects in the transcriptomic data, we calculated Probabilistic Estimation of Expression Residuals (PEER) factors. A covariate matrix was constructed including the first two genotype principal components (PCs), the first five PEER factors, age and sequencing platform. *FastQTL*^30^ was executed in nominal mode (without permutation) to compute cis-QTL associations for all variants within the 1-Mb cis-windows. Generating the full spectrum of QTLs across the loci, rather than retaining solely the most significant ones, was a strict prerequisite for the downstream Bayesian colocalization analysis.

Colocalization was performed utilizing the Approximate Bayes Factor (ABF) method implemented in the R package *coloc*^31^. GWAS summary statistics and the cis-QTL associations alleles were harmonized. For each locus, *coloc* computed the posterior probabilities across five mutually exclusive hypotheses: no genetic association with either trait (H0); association with sepsis survival only (H1); association with the molecular trait only (H2); associations with both traits driven by distinct SNVs (H3); and associations with both traits driven by a single shared SNV (H4). To declare colocalization evidence, a threshold of (H4) ≥ 80% was used.

### Functional enrichment

Gene Set Enrichment Analysis (GSEA) was performed utilizing the R package *clusterProfiler*^32^. Initially, all variants were mapped to their nearest proximal gene utilizing *bedtools*^33^ and ranked based on the minimum P-value of their top-associated index variant. Enrichment analysis was conducted across the Gene Ontology (GO), Kyoto Encyclopedia of Genes and Genomes (KEGG) databases, and Reactome pathways using the *ReactomePA*^34^ R package. P-values were adjusted utilizing the Benjamini-Hochberg (BH) procedure to control the False Discovery Rate (FDR). The statistical significance of each enriched term, scaled by its BH-adjusted P-value, was visualized as a colorimetric gradient utilizing dotplots (Supp. Fig. 5). To evaluate the directionality of the effects within the enriched biological networks, the −*log*_10_(*HR*) derived from the Cox proportional hazards models was mapped as a continuous colorimetric variable across the resulting gene-concept networks (cnetplots)^32^.

## Resource availability

### Lead contact

Further information and requests for resources should be directed to and will be fulfilled by the lead contact, Henri-Jean Garchon.

### Data and code availability

All original code used to pre-process, genotype, filter, and annotate (WGS) data is publicly available on GitHub and permanently archived on Zenodo (DOI: 10.5281/zenodo.24280139).

Any additional information required to reanalyze the data reported in this paper is available from the lead contact upon request.

## Acknowledgements

Funding information: RHU RECORDS (France 2030 program, ANR-18-RHUS-0004), iRECORDS funded by ERA PerMed (JTC_2021), IHU SEPSIS (France 2030 program, ANR-23-IAHU-0004).

## Supplementary figure legends

**Supp. Figure 1.**
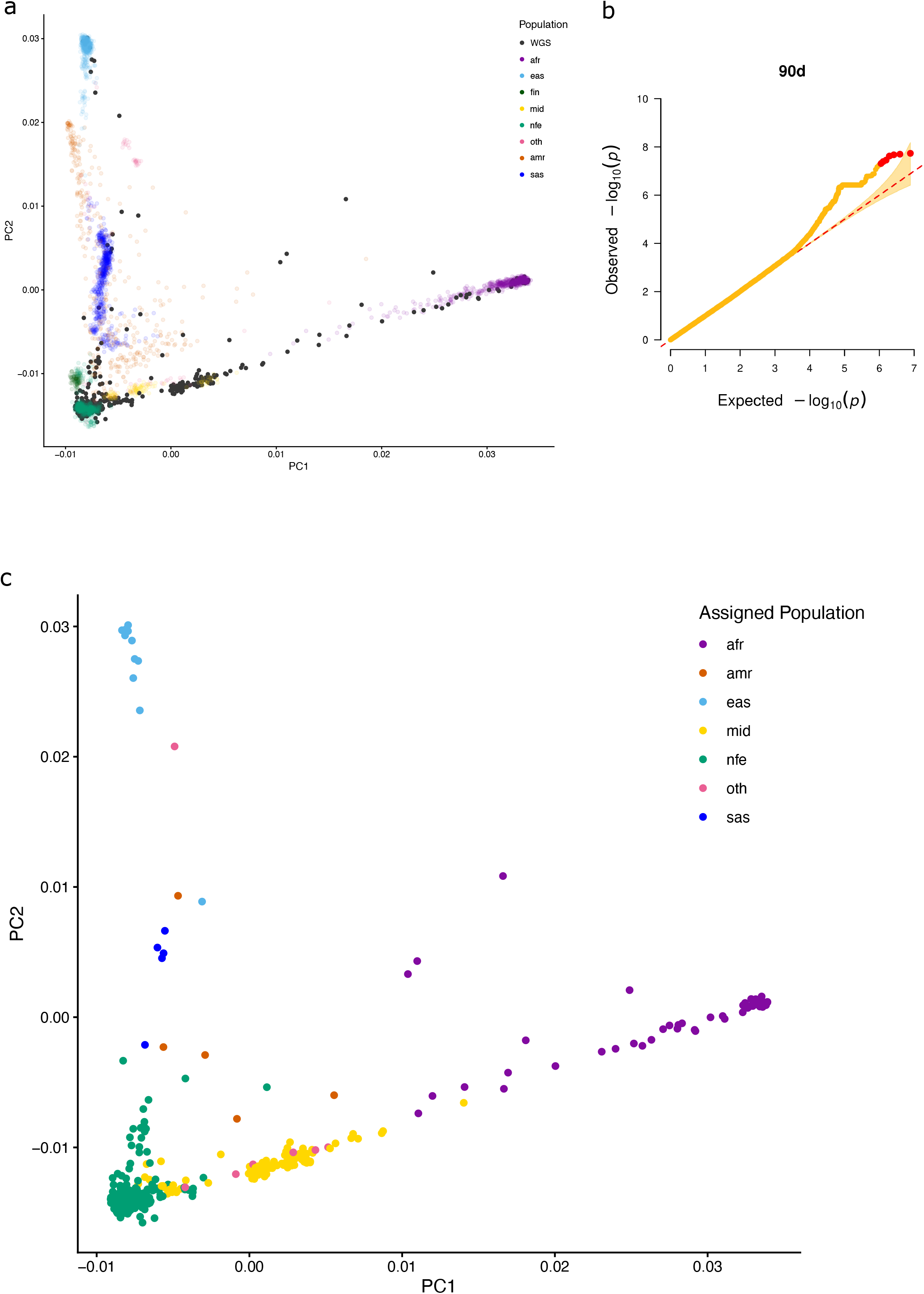
Principal component analysis and Q-Q plot of 90-day GWAS in sepsis patients. (a) Principal component analysis showing genetic ancestry in the study cohorts. Scatter plot displaying the first two principal components (PC1 on the x-axis and PC2 on the y-axis) to illustrate the population structure of the analyzed cohort. Patients from the study cohort (WGS, solid black dots) were projected onto the principal component space. Reference individuals are represented by circles and color-coded by their designated population ancestry group: African (afr, purple), Admixed American (amr, orange), East Asian (eas, light blue), Finnish (fin, dark green), Middle Eastern (mid, yellow), Non-Finnish European (nfe, teal), South Asian (sas, dark blue), and other/admixed ancestries (oth, pink). The projection demonstrates the distribution of genetic ancestries present within the sepsis patient cohort relative to known global populations. (b) Quantile-quantile (Q-Q) plot of genome-wide association study for late survival in sepsis patients. Q-Q plot comparing the observed (y-axis) against the expected (−log₁₀ P-values, x-axis) distribution for 90-day (orange dots) survival. The diagonal dashed red line represents the expected distribution of P-values under the null hypothesis of no genetic association. The shaded yellow region indicates the 95% confidence interval for the expected null distribution. The genomic inflation factor (λ) is λ= 1.00 for the 90-day analysis, indicating adequate control for population stratification and systemic bias. Red dots show p-values above the genome-wide significance threshold (5e-08). (c) Principal component analysis showing the assigned genetic ancestry of the sepsis samples. The projection demonstrates the distribution of genetic ancestries present within the sepsis patient cohort after (KNN) algorithm classification.

**Supp. Figure 2.**
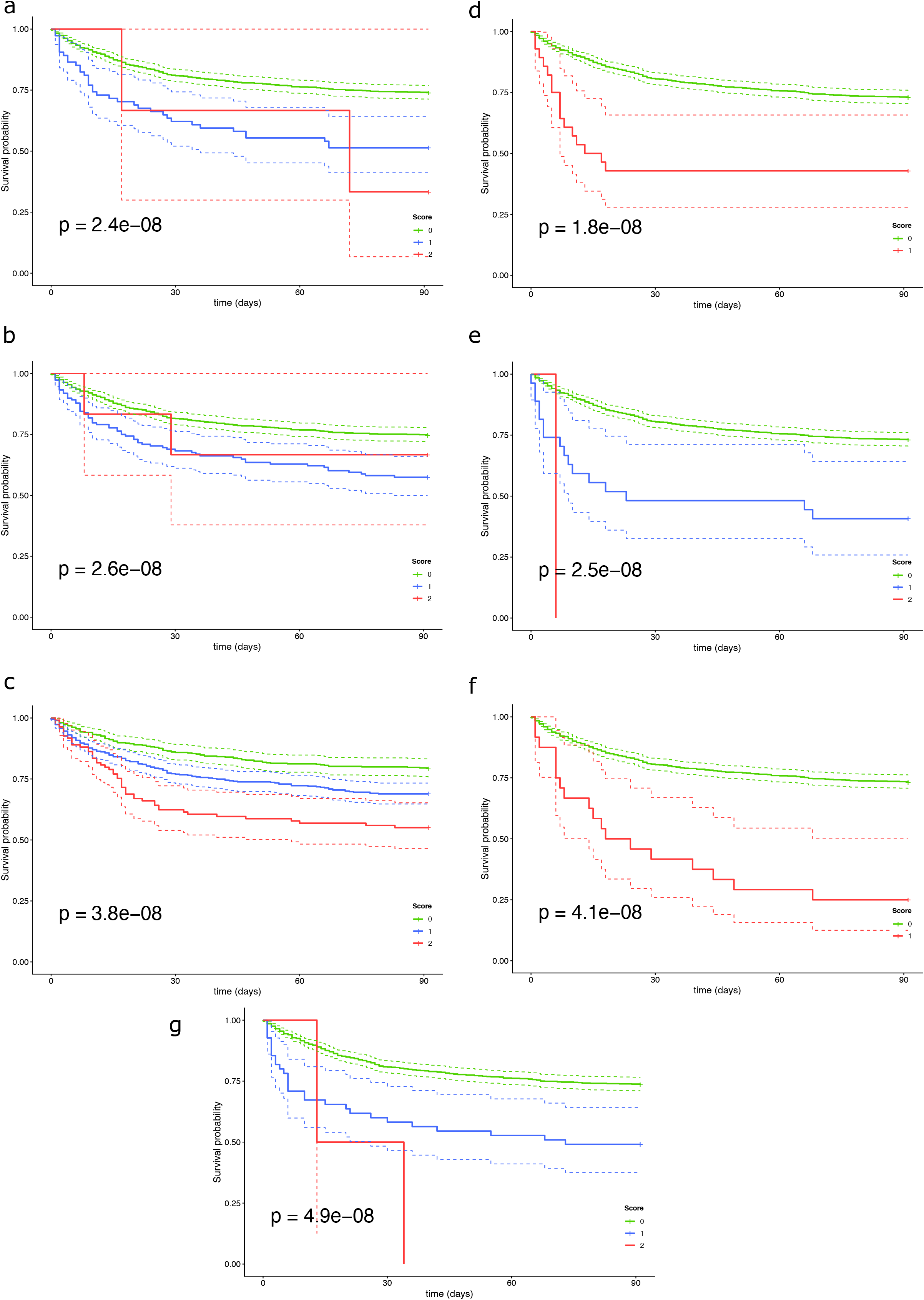
Kaplan-Meier survival curves for genome-wide significant variants associated with 90-day mortality in sepsis. (a-g) Kaplan-Meier estimates of 90-day survival probability for sepsis patients, stratified by genotype of top-associated variants at representative loci including *CALCRL-AS1, IMPDH1P7, PLCB1, ENSG00000288587, ENSG00000248172, ISG20, and RNU6-144P*, respectively. The x-axis represents time in days from study inclusion, and the y-axis indicates the estimated survival probability. Solid lines represent the survival trajectories for patient groups based on their effect allele dosage (Score 0 = homozygous reference, green; 1 = heterozygous, blue; 2 = homozygous alternative, red) under an additive genetic model. Dashed lines surrounding each curve denote the 95% confidence intervals. The P-values displayed in each panel reflect the statistical significance of the association derived from the corresponding Cox proportional hazards model.

**Supp. Figure 3.**
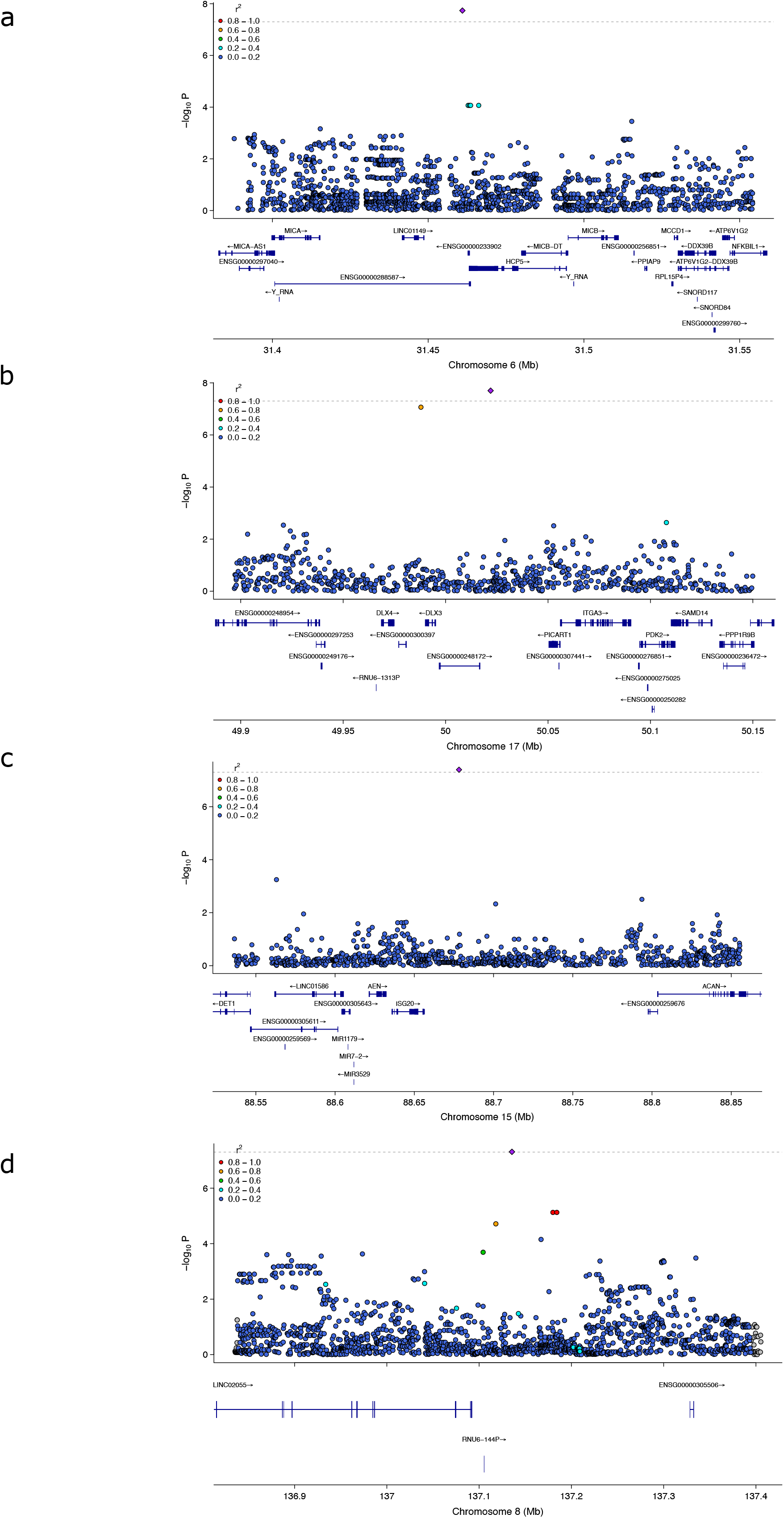
Regional association plots for genome-wide significant loci associated with 90-day survival in sepsis. (a-d) Locus zoom plots detailing the associated variants at loci including *ENSG00000288587, ENSG00000248172, ISG20, and RNU6-144P*, respectively. The top panel displays the association values −log₁₀(P) (y-axis) of SNVs according to their chromosomal position (x-axis). The top index variant at each locus is highlighted as a purple diamond. Surrounding variants are color-coded to reflect their linkage disequilibrium (r²) with the index variant (red, 0.8–1.0; orange, 0.6–0.8; green, 0.4–0.6; light blue, 0.2–0.4; dark blue, 0.0–0.2). The horizontal dashed grey line denotes the genome-wide significance threshold (P = 5 × 10⁻⁸). The bottom panels illustrate the local gene annotations and exon-intron structures, with arrows indicating the direction of transcription.

**Supp. Figure 4.**
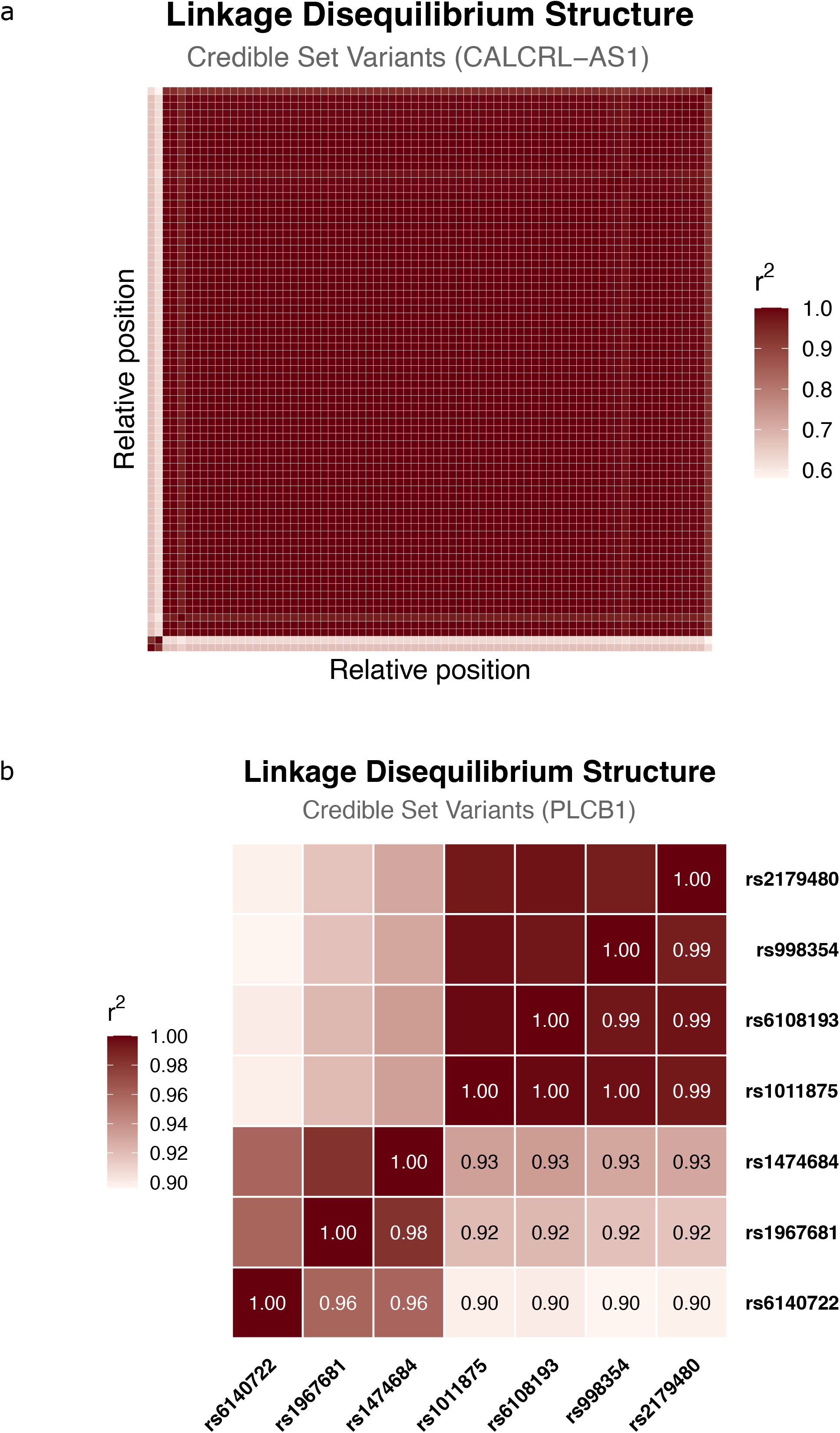
LD structure at credible sets of CALCRL-AS1 and PLCB1. (a) LD structure of the *CALCRL-AS1* credible set, including 75 variants. The *r*^2^ between every SNV in the LD matrix is color-coded with red. (b) LD structure of the *PLCB1* credible set, including 7 variants. SNVs are highlighted for each row, followed by their (*r*^2^).

**Supp. Figure 5.**
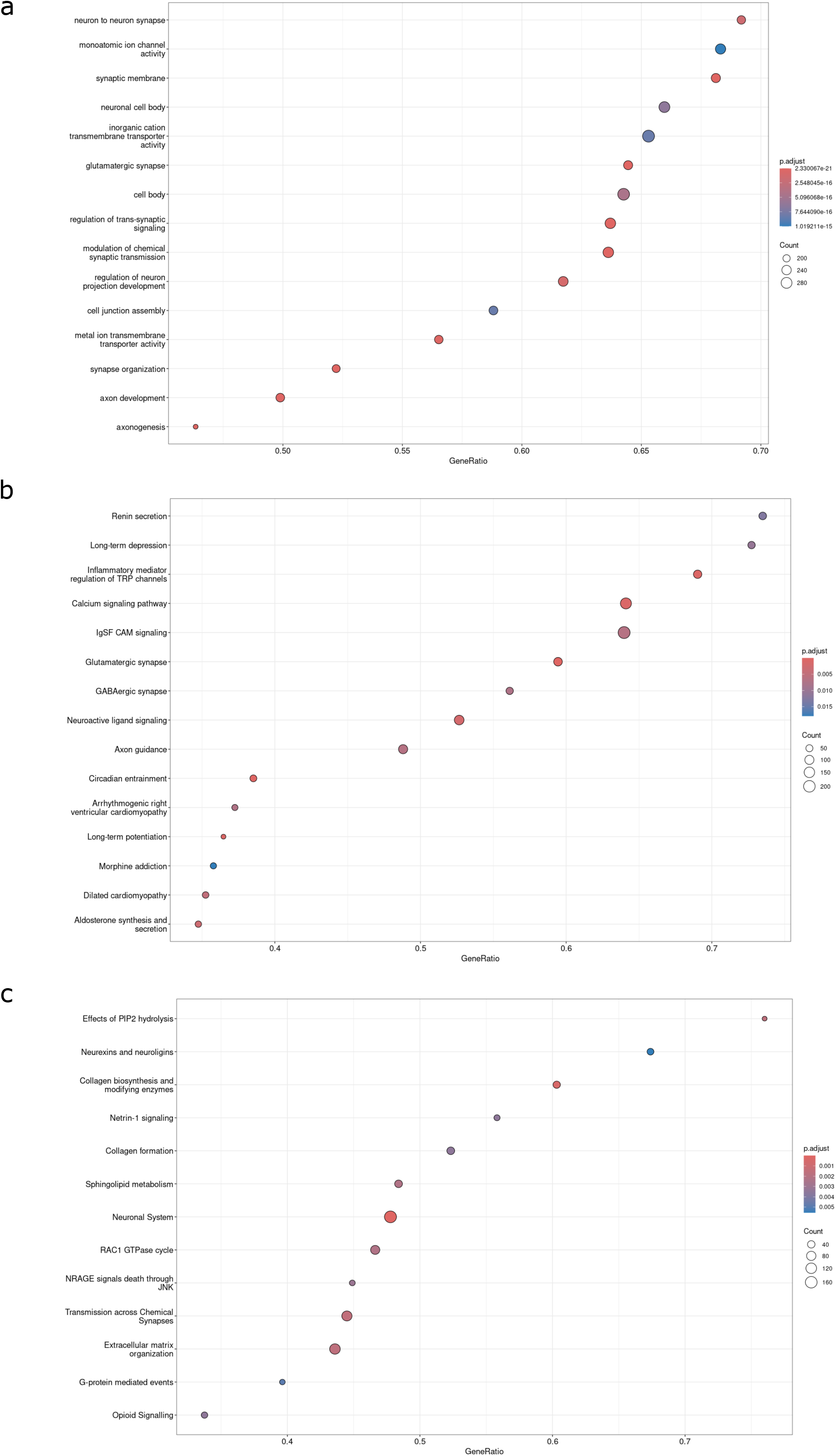
Dot-plots of enriched biological pathways associated with sepsis survival. Gene Set Enrichment Analysis (GSEA) networks relating pathways with genes across three annotation databases: Gene Ontology/GO (a), KEGG (b), and Reactome (c). Each row represents significantly enriched pathways or functional terms, with the size of the node proportional to the number of genes included in the specific category. Corresponding adjusted p-values of each term are color-coded based on their significance and illustrated in the circular nodes.

**Supp. Figure 6.**
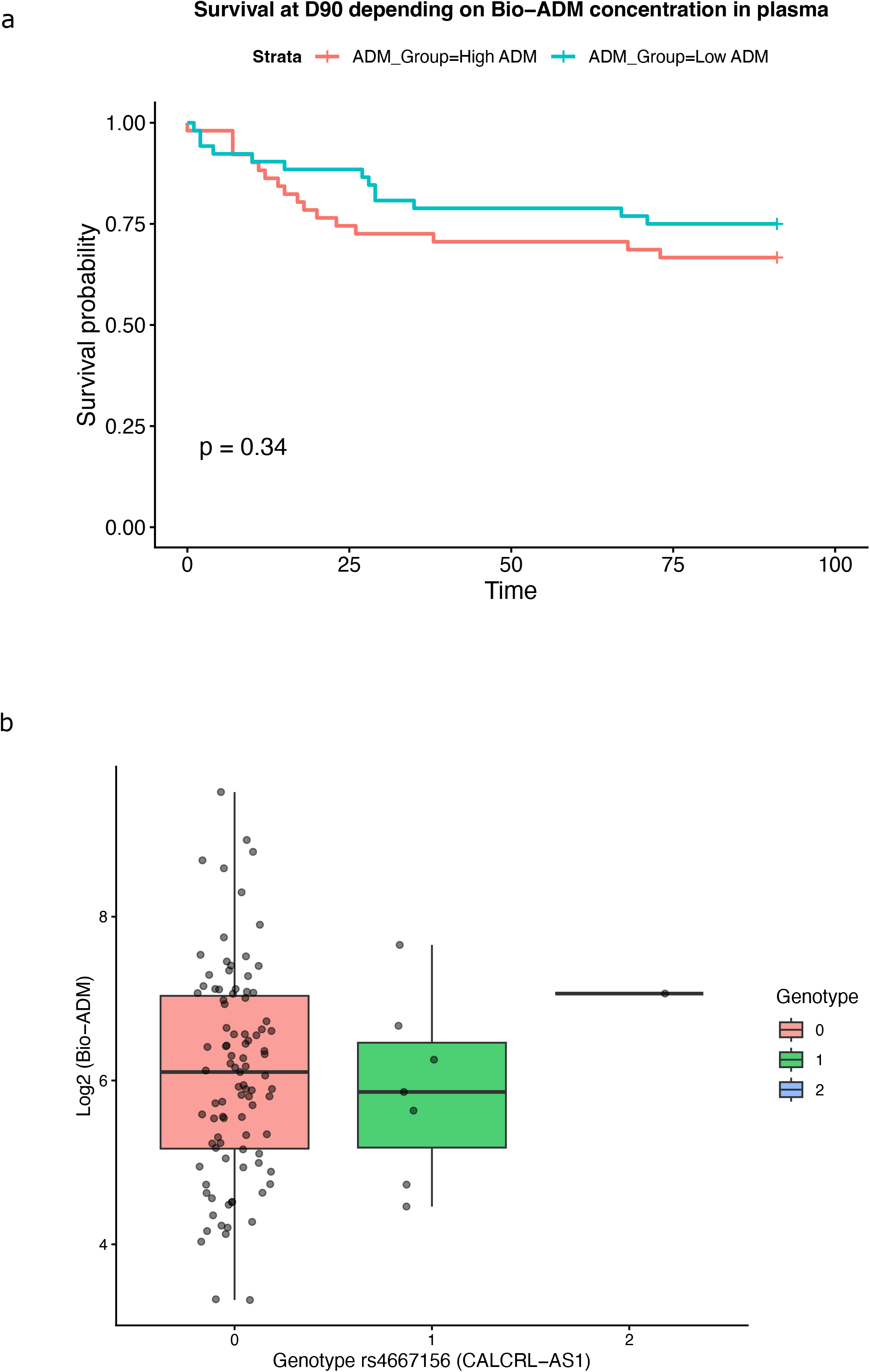
Analysis of circulating adrenomedullin (bio-ADM) in a subset of the TRIAL cohort. (a) Kaplan-Meier 90-day survival estimates stratified by plasma bio-ADM concentration. Patients were divided into high (above median) and low (below median) bio-ADM groups (log-rank P = 0.34). (b) Evaluation of the *CALCRL-AS1* index variant (rs4667156) on plasma bio-ADM concentrations. Log2-transformed bio-ADM levels are plotted across genotypes (0, 1, and 2) to test for a potential additive effect. No significant association was observed between genotype and circulating bio-ADM levels (Kruskal-Wallis *χ*^2^ = 0.94, P = 0.62).

**Supp. Figure 7.**
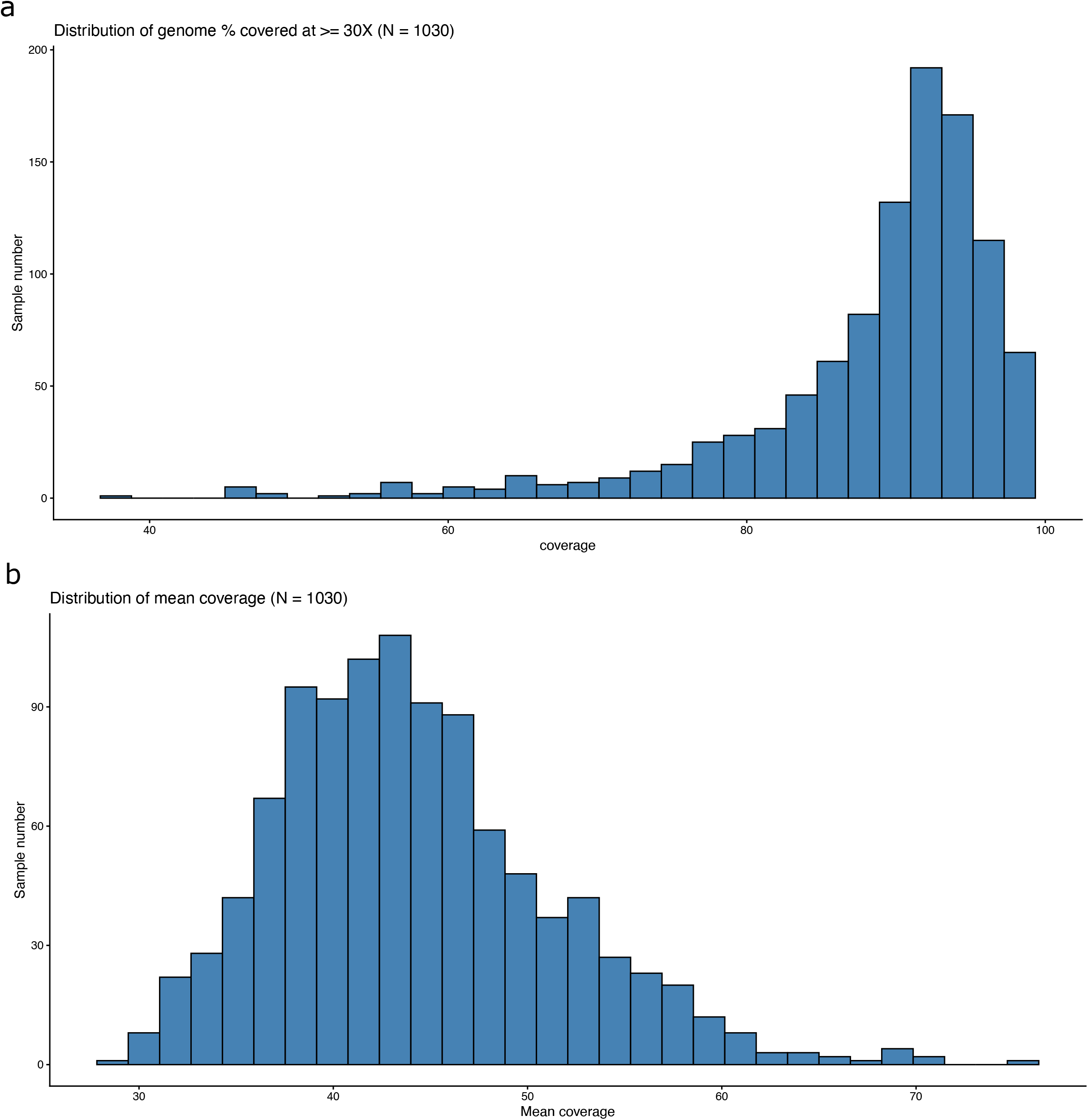
Histograms of sequencing depth and genome coverage. (a) Distribution of the genome percentage covered at ≥30X. (b) Distribution of the mean genome coverage.

**Supp. Figure 8.**
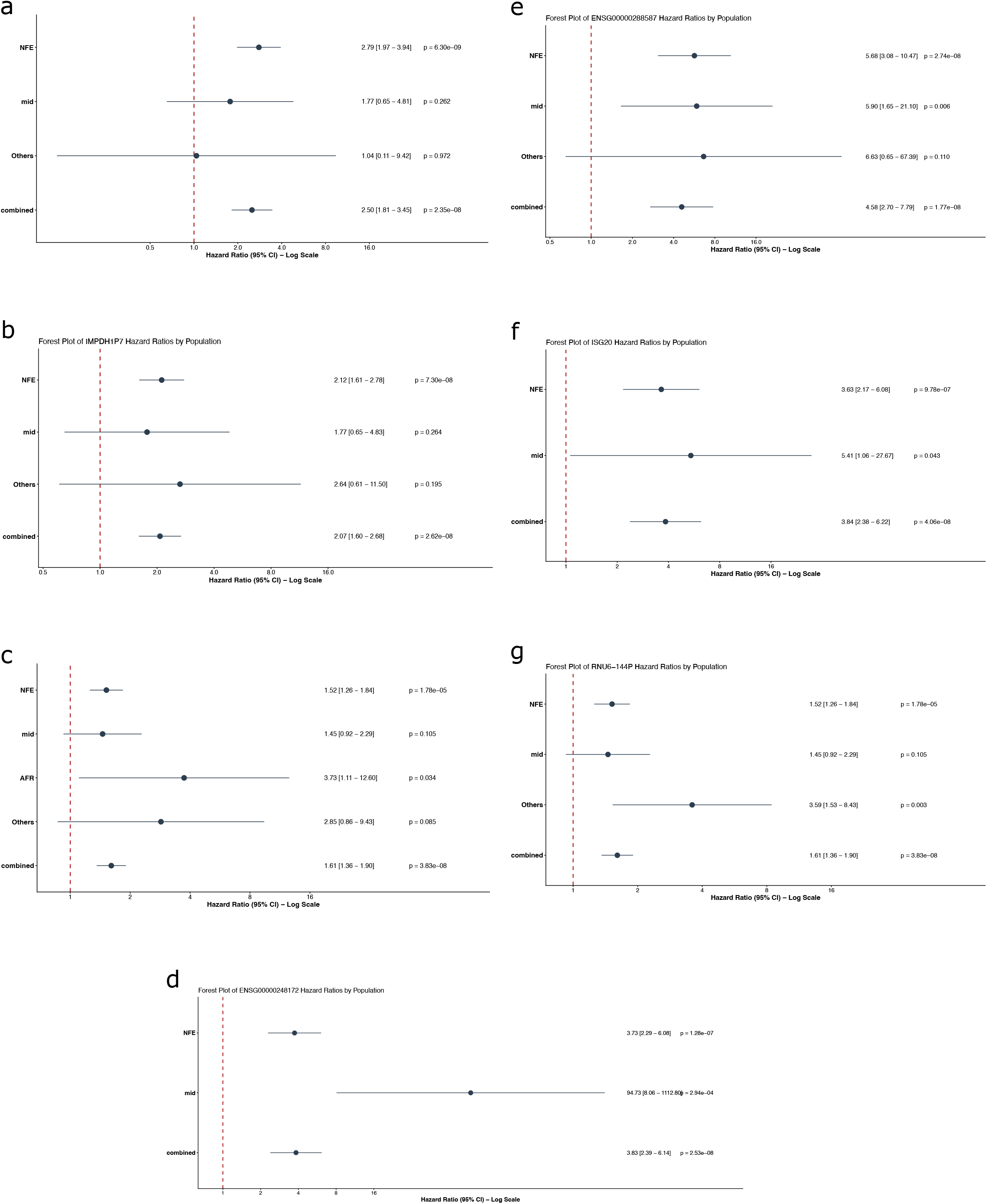
Forest plots of genome-wide significantly associated SNVs by population. (a-g) Population specific association P-values and HR of top-associated variants at representative loci including *CALCRL-AS1, IMPDH1P7, PLCB1, ENSG00000248172, ENSG00000288587, ISG20, and RNU6-144P,* respectively. Black circles represent the association p-value and HR (x-axis) with 95% confidence intervals (CI) for individual populations and the main analysis (y-axis). Horizontal dashed red line indicates the null effect (HR = 1). Solid vertical lines represent 95% CI.

## Supplementary tables

**Supplementary table 1.** Credible set of the *CALCRL-AS1* locus.

| SNP | POS | CHR | A1 | A0 | COEF | SE. COEF | PVALUE | PIP |
|---|---|---|---|---|---|---|---|---|
| rs146038228 | 186951556 | 2 | C | T | -7.31e-01 | 0.1309 | 2.38e-08 | 9.98e-02 |
| rs13032406 | 186959711 | 2 | T | G | -6.54e-01 | 0.1246 | 1.55e-07 | 1.92e-02 |
| rs73979608 | 187000629 | 2 | C | T | -8.86e-01 | 0.1745 | 3.80e-07 | 8.67e-03 |
| rs4667139 | 187005553 | 2 | A | T | -8.86e-01 | 0.1745 | 3.80e-07 | 8.67e-03 |
| rs55988918 | 187005848 | 2 | A | G | -8.87e-01 | 0.1693 | 1.61e-07 | 1.85e-02 |
| rs59938343 | 187010362 | 2 | A | G | -8.86e-01 | 0.1745 | 3.80e-07 | 8.67e-03 |
| rs55645636 | 187030901 | 2 | C | T | -8.86e-01 | 0.1745 | 3.80e-07 | 8.67e-03 |
| rs55895670 | 187034558 | 2 | C | T | -8.86e-01 | 0.1745 | 3.80e-07 | 8.67e-03 |
| rs73979617 | 187035811 | 2 | T | G | -8.86e-01 | 0.1745 | 3.80e-07 | 8.67e-03 |
| rs58465774 | 187041294 | 2 | A | T | -8.86e-01 | 0.1745 | 3.80e-07 | 8.67e-03 |
| rs57543473 | 187046289 | 2 | A | G | -8.86e-01 | 0.1745 | 3.80e-07 | 8.67e-03 |
| rs73979626 | 187057570 | 2 | A | T | -8.86e-01 | 0.1745 | 3.80e-07 | 8.67e-03 |
| rs73979628 | 187061868 | 2 | C | G | -8.86e-01 | 0.1745 | 3.80e-07 | 8.67e-03 |
| rs55763386 | 187065707 | 2 | T | C | -8.86e-01 | 0.1745 | 3.80e-07 | 8.67e-03 |
| rs73979641 | 187075471 | 2 | T | C | -8.86e-01 | 0.1745 | 3.80e-07 | 8.67e-03 |
| rs16828603 | 187077124 | 2 | C | T | -8.86e-01 | 0.1745 | 3.80e-07 | 8.67e-03 |
| rs75651380 | 187077675 | 2 | G | A | -8.86e-01 | 0.1745 | 3.80e-07 | 8.67e-03 |
| rs73979650 | 187089417 | 2 | G | T | -8.86e-01 | 0.1745 | 3.80e-07 | 8.67e-03 |
| rs73045608 | 187089569 | 2 | G | C | -8.86e-01 | 0.1745 | 3.80e-07 | 8.67e-03 |
| rs55670108 | 187094712 | 2 | G | C | -8.86e-01 | 0.1745 | 3.80e-07 | 8.67e-03 |
| rs10497679 | 187097095 | 2 | C | T | -8.86e-01 | 0.1745 | 3.80e-07 | 8.67e-03 |
| rs73979655 | 187098362 | 2 | G | A | -8.86e-01 | 0.1745 | 3.80e-07 | 8.67e-03 |
| rs73979656 | 187098375 | 2 | T | G | -8.86e-01 | 0.1745 | 3.80e-07 | 8.67e-03 |
| rs16828623 | 187104364 | 2 | C | T | -8.86e-01 | 0.1745 | 3.80e-07 | 8.67e-03 |
| rs79446017 | 187107537 | 2 | T | C | -8.86e-01 | 0.1745 | 3.80e-07 | 8.67e-03 |
| rs75622416 | 187107646 | 2 | C | G | -8.86e-01 | 0.1745 | 3.80e-07 | 8.67e-03 |
| rs74374012 | 187113747 | 2 | C | T | -8.86e-01 | 0.1745 | 3.80e-07 | 8.67e-03 |
| rs58223779 | 187118274 | 2 | A | G | -8.86e-01 | 0.1745 | 3.80e-07 | 8.67e-03 |
| rs73045652 | 187119075 | 2 | A | G | -8.86e-01 | 0.1745 | 3.80e-07 | 8.67e-03 |
| rs16828655 | 187128877 | 2 | A | T | -8.86e-01 | 0.1745 | 3.80e-07 | 8.67e-03 |
| rs56331249 | 187129839 | 2 | A | T | -8.86e-01 | 0.1745 | 3.80e-07 | 8.67e-03 |
| rs16828665 | 187135860 | 2 | C | A | -8.86e-01 | 0.1745 | 3.80e-07 | 8.67e-03 |
| rs16828674 | 187137847 | 2 | T | A | -8.86e-01 | 0.1745 | 3.80e-07 | 8.67e-03 |
| rs16828678 | 187141723 | 2 | A | T | -8.86e-01 | 0.1745 | 3.80e-07 | 8.67e-03 |
| rs73979681 | 187143950 | 2 | A | G | -8.86e-01 | 0.1745 | 3.80e-07 | 8.67e-03 |
| rs55845347 | 187145809 | 2 | A | G | -8.86e-01 | 0.1745 | 3.80e-07 | 8.67e-03 |
| rs73979685 | 187147179 | 2 | G | A | -8.86e-01 | 0.1745 | 3.80e-07 | 8.67e-03 |
| rs73979690 | 187149503 | 2 | T | G | -8.86e-01 | 0.1745 | 3.80e-07 | 8.67e-03 |
| rs4552154 | 187153244 | 2 | T | A | -8.86e-01 | 0.1745 | 3.80e-07 | 8.67e-03 |
| rs73979691 | 187156181 | 2 | G | A | -8.86e-01 | 0.1745 | 3.80e-07 | 8.67e-03 |
| rs59567244 | 187161907 | 2 | T | C | -8.86e-01 | 0.1745 | 3.80e-07 | 8.67e-03 |
| rs57108324 | 187162219 | 2 | C | G | -8.86e-01 | 0.1745 | 3.80e-07 | 8.67e-03 |
| rs77724824 | 187163501 | 2 | C | T | -8.86e-01 | 0.1745 | 3.80e-07 | 8.67e-03 |
| rs56740282 | 187167965 | 2 | G | A | -8.86e-01 | 0.1745 | 3.80e-07 | 8.67e-03 |
| rs4381752 | 187168318 | 2 | C | T | -8.86e-01 | 0.1745 | 3.80e-07 | 8.67e-03 |
| rs4465740 | 187168444 | 2 | T | C | -8.86e-01 | 0.1745 | 3.80e-07 | 8.67e-03 |
| rs73979695 | 187170225 | 2 | C | T | -8.86e-01 | 0.1745 | 3.80e-07 | 8.67e-03 |
| rs149619427 | 187171061 | 2 | T | C | -8.86e-01 | 0.1745 | 3.80e-07 | 8.67e-03 |
| rs145565469 | 187183653 | 2 | G | A | -8.86e-01 | 0.1745 | 3.80e-07 | 8.67e-03 |
| rs57928137 | 187187143 | 2 | G | C | -8.86e-01 | 0.1745 | 3.80e-07 | 8.67e-03 |
| rs55941489 | 187188071 | 2 | C | T | -8.86e-01 | 0.1745 | 3.80e-07 | 8.67e-03 |
| rs4233793 | 187189786 | 2 | C | G | -8.86e-01 | 0.1745 | 3.80e-07 | 8.67e-03 |
| rs4555314 | 187196035 | 2 | C | T | -8.86e-01 | 0.1745 | 3.80e-07 | 8.67e-03 |
| rs73981103 | 187197808 | 2 | G | A | -8.86e-01 | 0.1745 | 3.80e-07 | 8.67e-03 |
| rs4667152 | 187201320 | 2 | C | T | -8.86e-01 | 0.1745 | 3.80e-07 | 8.67e-03 |
| rs4413122 | 187201800 | 2 | C | T | -8.86e-01 | 0.1745 | 3.80e-07 | 8.67e-03 |
| rs1918908 | 187207083 | 2 | A | C | -8.86e-01 | 0.1745 | 3.80e-07 | 8.67e-03 |
| rs56045439 | 187216088 | 2 | G | T | -8.86e-01 | 0.1745 | 3.80e-07 | 8.67e-03 |
| rs73981118 | 187219165 | 2 | C | T | -8.86e-01 | 0.1745 | 3.80e-07 | 8.67e-03 |
| rs73981120 | 187219914 | 2 | A | G | -8.86e-01 | 0.1745 | 3.80e-07 | 8.67e-03 |
| rs73981121 | 187220349 | 2 | A | G | -8.86e-01 | 0.1745 | 3.80e-07 | 8.67e-03 |
| rs55637608 | 187227272 | 2 | G | A | -8.91e-01 | 0.1701 | 1.63e-07 | 1.83e-02 |
| rs55887912 | 187229255 | 2 | C | T | -8.91e-01 | 0.1701 | 1.63e-07 | 1.83e-02 |
| rs4667156 | 187232732 | 2 | A | G | -9.19e-01 | 0.164 | 2.12e-08 | 1.11e-01 |
| rs16828711 | 187237304 | 2 | G | C | -8.86e-01 | 0.1745 | 3.80e-07 | 8.67e-03 |
| rs16828719 | 187240351 | 2 | C | T | -8.86e-01 | 0.1745 | 3.80e-07 | 8.67e-03 |
| rs73981127 | 187242229 | 2 | G | A | -8.86e-01 | 0.1745 | 3.80e-07 | 8.67e-03 |
| rs59832815 | 187243390 | 2 | G | T | -8.86e-01 | 0.1745 | 3.80e-07 | 8.67e-03 |
| rs79005508 | 187244028 | 2 | A | G | -8.86e-01 | 0.1745 | 3.80e-07 | 8.67e-03 |
| rs56235386 | 187244420 | 2 | G | A | -8.86e-01 | 0.1745 | 3.80e-07 | 8.67e-03 |
| rs73981133 | 187255293 | 2 | A | G | -8.93e-01 | 0.1744 | 3.01e-07 | 1.06e-02 |
| rs57732150 | 187260036 | 2 | A | G | -8.93e-01 | 0.1744 | 3.01e-07 | 1.06e-02 |
| rs144132496 | 187260700 | 2 | A | C | -8.93e-01 | 0.1744 | 3.01e-07 | 1.06e-02 |
| rs73981136 | 187263970 | 2 | G | A | -8.93e-01 | 0.1744 | 3.01e-07 | 1.06e-02 |
| rs74865511 | 187378134 | 2 | A | G | -8.86e-01 | 0.1692 | 1.63e-07 | 1.83e-02 |

**Supplementary table 2.** Credible set of the *PLCB1* locus.

| SNP | POS | CHR | A1 | A0 | COEF | SE. COEF | PVALUE | PIP |
|---|---|---|---|---|---|---|---|---|
| rs6140722 | 8795463 | 20 | C | T | -4.41e-01 | 0.08685 | 3.78e-07 | 7.48e-02 |
| rs1967681 | 8795683 | 20 | A | G | -4.56e-01 | 0.0864 | 1.34e-07 | 1.89e-01 |
| rs1474684 | 8795913 | 20 | A | G | -4.76e-01 | 0.08632 | 3.51e-08 | 6.19e-01 |
| rs1011875 | 8797442 | 20 | A | C | -4.03e-01 | 0.08559 | 2.44e-06 | 1.51e-02 |
| rs6108193 | 8797599 | 20 | A | T | -4.09e-01 | 0.08546 | 1.69e-06 | 2.06e-02 |
| rs998354 | 8798100 | 20 | T | G | -4.04e-01 | 0.08562 | 2.34e-06 | 1.57e-02 |
| rs2179480 | 8798196 | 20 | T | C | -4.19e-01 | 0.08542 | 9.29e-07 | 3.46e-02 |

Supplementary table 3 List of eQTLs around *CALCRL-AS1* locus, available at DOI: 10.5281/zenodo.24280139

Supplementary table 4 List of eQTLs around *PLCB1* locus, available at DOI: 10.5281/zenodo.24280139

Supplementary table 5 List of sQTLs around *CALCRL-AS1* locus, available at DOI: 10.5281/zenodo.24280139

Supplementary table 6 List of sQTLs around *PLCB1* locus, available at DOI: 10.5281/zenodo.24280139

**Supplementary table 7.** Colocalization with eQTLs at the *CALCRL-AS1* locus.

| Gene | HGNC symbol | H0 | H1 | H2 | H3 | H4 | Common SNVs |
|---|---|---|---|---|---|---|---|
| ENSG00000259915 |  | 3.37 | 48.4 | 1.29 | 18.46 | 28.47 | 1812 |
| ENSG00000237877 | *LINC01473* | 9.69 | 55.45 | 1.49 | 8.51 | 24.86 | 873 |
| ENSG00000144369 | *FAM171B* | 4.32 | 64.39 | 1.22 | 18.14 | 11.93 | 2574 |
| ENSG00000003436 | *TFPI* | 4.57 | 68.22 | 1.1 | 16.47 | 9.63 | 2631 |
| ENSG00000064989 | *CALCRL* | 4.54 | 67.67 | 1.3 | 19.4 | 7.09 | 2631 |
| ENSG00000138448 | *ITGAV* | 4.74 | 70.35 | 1.14 | 16.9 | 6.86 | 2347 |
| ENSG00000065548 | *ZC3H15* | 5.08 | 74.89 | 0.9 | 13.29 | 5.84 | 2111 |
| ENSG00000144366 | *GULP1* | 49.84 | 38.31 | 4.37 | 3.35 | 4.13 | 1046 |
| ENSG00000287129 |  | 88.37 | 5.86 | 4.91 | 0.33 | 0.53 | 645 |
| ENSG00000188738 | *FSIP2* | 99.83 | 0.08 | 0.08 | 0.00e+00 | 0.01 | 10 |

**Supplementary table 8.** Colocalization with sQTLs at the *CALCRL-AS1* locus.

| Gene | Phenotype ID | H0 | H1 | H2 | H3 | H4 | Common SNVs |
|---|---|---|---|---|---|---|---|
| *CALCRL* | chr2:187520619:187554200:clu_11496_NA | 4.73 | 70.53 | 1.04 | 15.55 | 8.14 | 2631 |
| *CALCRL* | chr2:187431365:187436598:clu_11495_NA | 4.59 | 68.5 | 1.2 | 17.85 | 7.86 | 2631 |
| *CALCRL* | chr2:187522888:187554200:clu_11496_NA | 4.75 | 70.8 | 1.05 | 15.59 | 7.83 | 2631 |
| *CALCRL* | chr2:187520619:187522624:clu_11496_NA | 4.73 | 70.55 | 1.07 | 15.88 | 7.77 | 2631 |
| *CALCRL* | chr2:187484988:187496881:clu_11497_NA | 4.7 | 70.04 | 1.1 | 16.45 | 7.71 | 2631 |
| *CALCRL* | chr2:187513647:187529364:clu_11496_NA | 4.72 | 70.47 | 1.08 | 16.08 | 7.64 | 2631 |
| *CALCRL* | chr2:187503771:187513616:clu_11496_NA | 4.6 | 68.57 | 1.21 | 18.06 | 7.56 | 2631 |
| *CALCRL* | chr2:187503771:187554200:clu_11496_NA | 4.34 | 64.72 | 1.48 | 22.03 | 7.43 | 2631 |
| *CALCRL* | chr2:187513647:187554200:clu_11496_NA | 4.71 | 70.31 | 1.1 | 16.46 | 7.41 | 2631 |
| *CALCRL* | chr2:187513647:187520542:clu_11496_NA | 4.75 | 70.8 | 1.07 | 16 | 7.38 | 2631 |
| *CALCRL* | chr2:187549515:187554200:clu_11496_NA | 4.77 | 71.15 | 1.06 | 15.73 | 7.29 | 2631 |
| *CALCRL* | chr2:187544638:187554200:clu_11496_NA | 4.77 | 71.22 | 1.05 | 15.69 | 7.26 | 2631 |
| *CALCRL* | chr2:187467933:187554200:clu_11496_NA | 4.79 | 71.47 | 1.05 | 15.68 | 7 | 2631 |
| *CALCRL* | chr2:187387757:187431297:clu_11495_NA | 4.79 | 71.44 | 1.06 | 15.82 | 6.9 | 2631 |
| *CALCRL* | chr2:187522882:187554200:clu_11496_NA | 4.81 | 71.81 | 1.04 | 15.48 | 6.86 | 2631 |
| *CALCRL* | chr2:187522864:187554200:clu_11496_NA | 4.63 | 69.13 | 1.22 | 18.17 | 6.86 | 2631 |
| *CALCRL* | chr2:187503771:187528701:clu_11496_NA | 4.83 | 72 | 1.03 | 15.33 | 6.81 | 2631 |
| *CALCRL* | chr2:187503771:187520542:clu_11496_NA | 4.77 | 71.13 | 1.09 | 16.25 | 6.75 | 2631 |
| *CALCRL* | chr2:187549515:187550839:clu_11496_NA | 4.76 | 71.04 | 1.1 | 16.4 | 6.7 | 2631 |
| *CALCRL* | chr2:187520619:187529364:clu_11496_NA | 4.8 | 71.55 | 1.07 | 15.89 | 6.69 | 2631 |
| *CALCRL* | chr2:187503771:187550839:clu_11496_NA | 4.77 | 71.13 | 1.1 | 16.39 | 6.62 | 2631 |
| *CALCRL* | chr2:187529486:187554200:clu_11496_NA | 4.63 | 69.03 | 1.24 | 18.49 | 6.62 | 2631 |
| *CALCRL* | chr2:187520614:187529364:clu_11496_NA | 4.8 | 71.66 | 1.06 | 15.85 | 6.62 | 2631 |
| *CALCRL* | chr2:187520614:187554200:clu_11496_NA | 4.81 | 71.75 | 1.06 | 15.86 | 6.52 | 2631 |
| *CALCRL* | chr2:187478804:187484124:clu_11496_NA | 4.8 | 71.6 | 1.08 | 16.05 | 6.47 | 2631 |
| *CALCRL* | chr2:187503771:187529364:clu_11496_NA | 4.76 | 70.99 | 1.12 | 16.68 | 6.46 | 2631 |
| *CALCRL* | chr2:187467933:187484124:clu_11496_NA | 4.74 | 70.75 | 1.14 | 16.93 | 6.43 | 2631 |
| *CALCRL* | chr2:187545104:187554200:clu_11496_NA | 4.77 | 71.14 | 1.11 | 16.58 | 6.4 | 2631 |
| *CALCRL* | chr2:187522864:187529364:clu_11496_NA | 4.82 | 71.86 | 1.07 | 15.97 | 6.28 | 2631 |
| *CALCRL* | chr2:187529486:187549460:clu_11496_NA | 4.85 | 72.32 | 1.04 | 15.58 | 6.21 | 2631 |
| *CALCRL* | chr2:187428890:187431297:clu_11495_NA | 4.39 | 65.56 | 1.5 | 22.37 | 6.18 | 2631 |
| *CALCRL* | chr2:187503771:187544504:clu_11496_NA | 4.87 | 72.62 | 1.03 | 15.32 | 6.15 | 2631 |
| *CALCRL* | chr2:187436657:187448039:clu_11495_NA | 4.62 | 68.95 | 1.28 | 19.03 | 6.13 | 2631 |
| *CALCRL* | chr2:187503771:187522624:clu_11496_NA | 4.74 | 70.71 | 1.16 | 17.33 | 6.05 | 2631 |
| *CALCRL* | chr2:187484988:187487806:clu_11497_NA | 4.72 | 70.4 | 1.19 | 17.72 | 5.97 | 2631 |
| *CALCRL* | chr2:187442039:187448039:clu_11495_NA | 4.61 | 68.82 | 1.3 | 19.39 | 5.87 | 2631 |
| *CALCRL* | chr2:187484988:187488337:clu_11497_NA | 4.77 | 71.08 | 1.15 | 17.17 | 5.84 | 2631 |
| *CALCRL* | chr2:187387757:187441923:clu_11495_NA | 4.57 | 68.15 | 1.36 | 20.23 | 5.7 | 2631 |
| *CALCRL* | chr2:187436657:187441923:clu_11495_NA | 4.65 | 69.37 | 1.28 | 19.04 | 5.67 | 2631 |
| *CALCRL* | chr2:187387757:187436598:clu_11495_NA | 4.77 | 71.14 | 1.16 | 17.33 | 5.6 | 2631 |
| *CALCRL* | chr2:187488376:187496881:clu_11497_NA | 4.9 | 73.1 | 1.03 | 15.39 | 5.59 | 2631 |
| *CALCRL* | chr2:187387466:187387666:clu_11494_NA | 4.54 | 67.78 | 1.39 | 20.71 | 5.57 | 2631 |
| *CALCRL* | chr2:187487874:187488337:clu_11497_NA | 4.67 | 69.73 | 1.26 | 18.81 | 5.52 | 2631 |
| *CALCRL* | chr2:187385632:187387329:clu_11494_NA | 4.71 | 70.3 | 1.22 | 18.26 | 5.51 | 2631 |
| *CALCRL* | chr2:187431365:187448039:clu_11495_NA | 4.76 | 71.01 | 1.18 | 17.55 | 5.5 | 2631 |
| *CALCRL* | chr2:187385632:187387666:clu_11494_NA | 4.79 | 71.47 | 1.15 | 17.12 | 5.47 | 2631 |
| *CALCRL* | chr2:187387757:187448039:clu_11495_NA | 4.65 | 69.37 | 1.3 | 19.32 | 5.37 | 2631 |
| *CALCRL* | chr2:187387494:187387666:clu_11494_NA | 4.62 | 68.97 | 1.33 | 19.76 | 5.32 | 2631 |

**Supplementary table 9.**
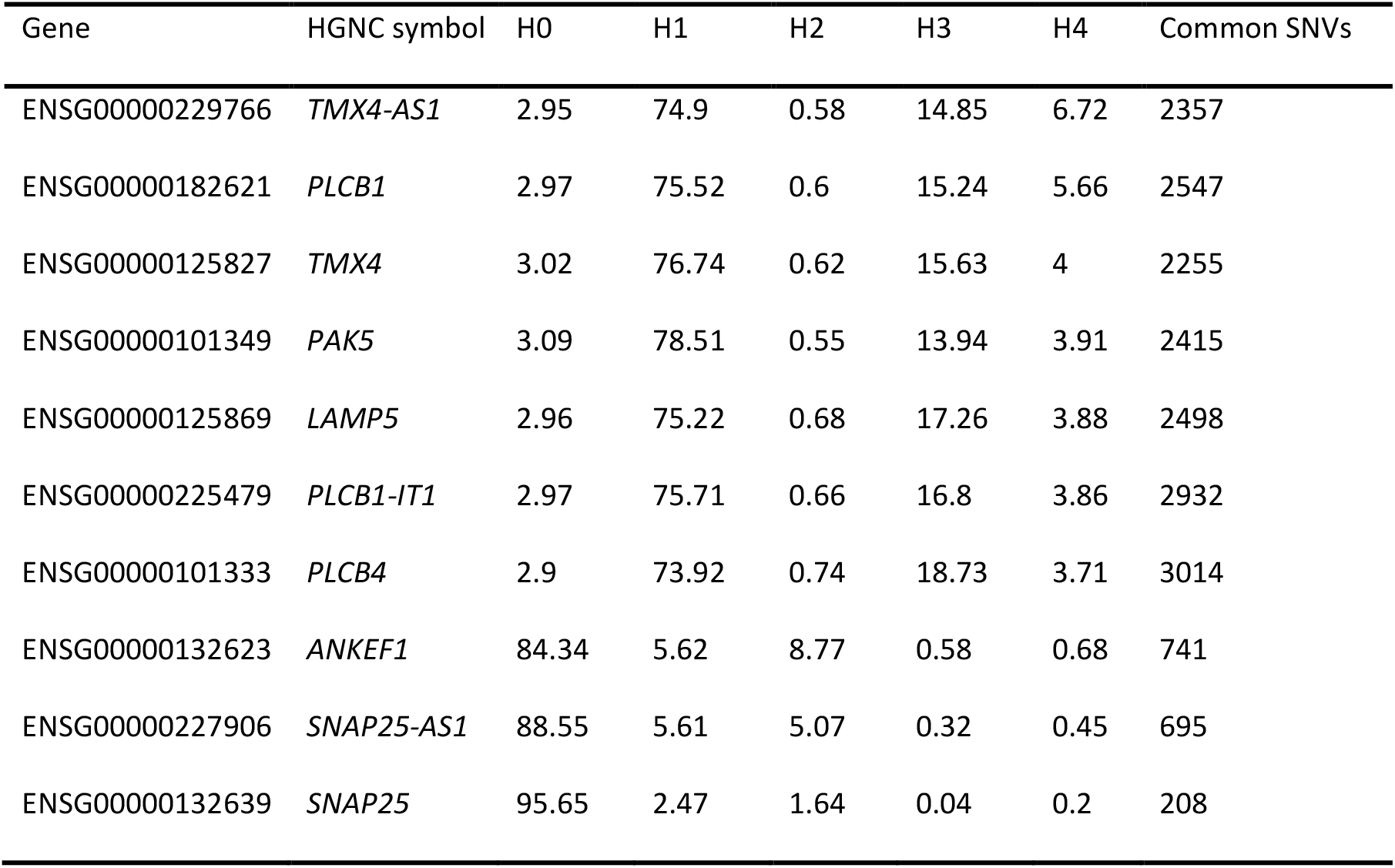
Colocalization with eQTLs at the *PLCB1* locus.

| Gene | HGNC symbol | H0 | H1 | H2 | H3 | H4 | Common SNVs |
|---|---|---|---|---|---|---|---|
| ENSG00000229766 | *TMX4-AS1* | 2.95 | 74.9 | 0.58 | 14.85 | 6.72 | 2357 |
| ENSG00000182621 | *PLCB1* | 2.97 | 75.52 | 0.6 | 15.24 | 5.66 | 2547 |
| ENSG00000125827 | *TMX4* | 3.02 | 76.74 | 0.62 | 15.63 | 4 | 2255 |
| ENSG00000101349 | *PAK5* | 3.09 | 78.51 | 0.55 | 13.94 | 3.91 | 2415 |
| ENSG00000125869 | *LAMP5* | 2.96 | 75.22 | 0.68 | 17.26 | 3.88 | 2498 |
| ENSG00000225479 | *PLCB1-IT1* | 2.97 | 75.71 | 0.66 | 16.8 | 3.86 | 2932 |
| ENSG00000101333 | *PLCB4* | 2.9 | 73.92 | 0.74 | 18.73 | 3.71 | 3014 |
| ENSG00000132623 | *ANKEF1* | 84.34 | 5.62 | 8.77 | 0.58 | 0.68 | 741 |
| ENSG00000227906 | *SNAP25-AS1* | 88.55 | 5.61 | 5.07 | 0.32 | 0.45 | 695 |
| ENSG00000132639 | *SNAP25* | 95.65 | 2.47 | 1.64 | 0.04 | 0.2 | 208 |

**Supplementary table 10.** Colocalization with sQTLs at the PLCB1 locus.

| Gene | Phenotype ID | H0 | H1 | H2 | H3 | H4 | Common SNVs |
|---|---|---|---|---|---|---|---|
| *PLCB1* | chr20:8732171:8733238:clu_69312_NA | 1.73 | 44.13 | 0.5 | 12.73 | 40.91 | 3014 |
| *PLCB1* | chr20:8404481:8406282:clu_69305_NA | 2.53 | 64.54 | 0.65 | 16.64 | 15.64 | 3014 |
| *PLCB1* | chr20:8399168:8406282:clu_69305_NA | 2.55 | 64.91 | 0.69 | 17.52 | 14.34 | 3014 |
| *PLCB1* | chr20:8324131:8371382:clu_69304_NA | 2.6 | 66.17 | 0.69 | 17.45 | 13.1 | 3014 |
| *PLCB1* | chr20:8371451:8399099:clu_69305_NA | 2.65 | 67.46 | 0.69 | 17.6 | 11.6 | 3014 |
| *PLCB1* | chr20:8371451:8406282:clu_69305_NA | 2.7 | 68.91 | 0.76 | 19.24 | 8.4 | 3014 |
| *PLCB1* | chr20:8649450:8651413:clu_69311_NA | 2.78 | 70.75 | 0.73 | 18.72 | 7.02 | 3014 |
| *PLCB1* | chr20:8741574:8750804:clu_69314_NA | 2.8 | 71.39 | 0.72 | 18.26 | 6.83 | 3014 |
| *PLCB1* | chr20:8729175:8733238:clu_69312_NA | 2.81 | 71.63 | 0.73 | 18.51 | 6.32 | 3014 |
| *PLCB1* | chr20:8750916:8751122:clu_69314_NA | 2.82 | 71.98 | 0.74 | 18.74 | 5.72 | 3014 |
| *PLCB1* | chr20:8305934:8371382:clu_69304_NA | 2.83 | 72.05 | 0.74 | 18.82 | 5.57 | 3014 |
| *PLCB1* | chr20:8150372:8371382:clu_69304_NA | 2.65 | 67.48 | 0.92 | 23.48 | 5.47 | 2760 |
| *PLCB1* | chr20:8207469:8371382:clu_69304_NA | 2.88 | 73.26 | 0.7 | 17.77 | 5.39 | 2877 |
| PLCB1 | chr20:8657285:8658538:clu_69311_NA | 2.82 | 71.88 | 0.76 | 19.33 | 5.21 | 3014 |
| *PLCB1* | chr20:8739361:8740344:clu_69313_NA | 2.75 | 70 | 0.84 | 21.42 | 4.99 | 3014 |
| *PLCB1* | chr20:8750916:8757046:clu_69314_NA | 2.85 | 72.58 | 0.74 | 18.93 | 4.9 | 3014 |
| *PLCB1* | chr20:8790262:8792085:clu_69315_NA | 2.88 | 73.42 | 0.72 | 18.33 | 4.65 | 3014 |
| *PLCB1* | chr20:8600782:8628294:clu_69305_NA | 2.85 | 72.71 | 0.75 | 19.08 | 4.61 | 3014 |
| *PLCB1* | chr20:8150384:8371382:clu_69304_NA | 2.89 | 73.49 | 0.72 | 18.31 | 4.6 | 2760 |
| *PLCB1* | chr20:8371451:8628294:clu_69305_NA | 2.85 | 72.74 | 0.75 | 19.08 | 4.57 | 3014 |
| *PLCB1* | chr20:8649450:8657184:clu_69311_NA | 2.89 | 73.6 | 0.73 | 18.48 | 4.3 | 3014 |
| *PLCB1* | chr20:8790262:8802056:clu_69315_NA | 2.89 | 73.56 | 0.73 | 18.7 | 4.12 | 3014 |
| *PLCB1* | chr20:8649450:8651628:clu_69311_NA | 2.88 | 73.42 | 0.74 | 18.93 | 4.02 | 3014 |
| *PLCB1* | chr20:8802174:8803839:clu_69315_NA | 2.84 | 72.35 | 0.79 | 20.02 | 4.01 | 3014 |
| *PLCB1* | chr20:8802174:8854683:clu_69315_NA | 2.85 | 72.53 | 0.78 | 19.88 | 3.97 | 3014 |
| *PLCB1* | chr20:8649450:8658538:clu_69311_NA | 2.9 | 73.94 | 0.73 | 18.5 | 3.94 | 3014 |
| *PLCB1* | chr20:8739361:8740277:clu_69313_NA | 2.85 | 72.71 | 0.77 | 19.74 | 3.92 | 3014 |
| *PLCB1* | chr20:8741574:8757046:clu_69314_NA | 2.89 | 73.55 | 0.74 | 18.95 | 3.87 | 3014 |
| *PLCB1* | chr20:8802174:8962196:clu_69315_NA | 2.86 | 72.93 | 0.77 | 19.6 | 3.84 | 3014 |
| *PLCB1* | chr20:8647954:8649397:clu_69310_NA | 2.78 | 70.78 | 0.85 | 21.77 | 3.82 | 3014 |
| *PLCB1* | chr20:8647954:8649374:clu_69310_NA | 2.89 | 73.53 | 0.75 | 19.02 | 3.82 | 3014 |
| *PLCB1* | chr20:8802174:8881622:clu_69315_NA | 2.77 | 70.64 | 0.86 | 22 | 3.72 | 3014 |
| *PLCB1* | chr20:8647954:8649394:clu_69310_NA | 2.69 | 68.54 | 0.95 | 24.17 | 3.65 | 3014 |
| *PLCB1* | chr20:8790262:8881622:clu_69315_NA | 2.75 | 70.13 | 0.89 | 22.6 | 3.64 | 3014 |

**Supplementary table 11.** Replication of previously reported associations in our 90-day survival analysis.

| First Author | Tested phenotype | Significant associations | Gene | Discovery P-value | Population MAF | Sample MAF | Replication P-value |
|---|---|---|---|---|---|---|---|
| D'urso et al. | Case vs Control | 1 | *AL589740.1* | 1.05e-10 | 0.170 | 0.129 | 0.52 |
| Guillen-Guio et al. | Case vs Control | 1 | *FLT1* | 5.18e-08 | 0.250 | 0.288 | 0.14 |
| Hernandez-Beeftink et al. | 28-day Survival | 3 | *SAMD9* | 4.9e-08 | 0.006 | / | / |
|  |  |  | *SLC5A12* | 3.0e-08 | 0.026 | 0.013 | 0.34 |
|  |  |  | *LINC00378* | 4.4e-08 | 0.008 | 0.012 | 0.61 |
| Rautanen et al. | 28-day Survival | 1 | *FER* | 5.6e-08 | 0.187 | 0.179 | 0.39 |

**Supplementary table 12.** Distribution of sequencing depth and genome coverage.

| Metric | Min | 1<sup>st</sup> Qu. | Median | Mean | 3<sup>rd</sup> Qu. | Max |
|---|---|---|---|---|---|---|
| Mean coverage | 28.64 | 39.02 | 43.30 | 44.17 | 47.82 | 75.51 |
| % of genome covered ≥ 10X | 97.16 | 98.67 | 99.22 | 99.07 | 99.37 | 99.58 |
| % of genome covered ≥ 30X | 37.74 | 86.25 | 91.27 | 88.65 | 94.23 | 98.29 |
| % of duplicates | 4.73 | 7.31 | 8.61 | 9.01 | 9.91 | 23.49s |

**Supplementary table 13.** Average sequencing depth of significantly associated variants. Chromosome.

| Chromosome | Position | rsID | Ref | Alt | Total DP | DP ($\frac{Total\ DP}{1065}$) |
|---|---|---|---|---|---|---|
| chr6 | 31460992 | rs11754929 | C | T | 41694 | 39.149 |
| chr17 | 50021979 | rs74570843 | C | T | 40776 | 38.287 |
| chr2 | 187232732 | rs4667156 | A | G | 39763 | 37.336 |
| chr2 | 186951556 | rs146038228 | C | T | 43992 | 41.307 |
| chr20 | 8795913 | rs1474684 | G | A | 42641 | 40.038 |
| chr15 | 88678201 | rs139793868 | C | G | 40449 | 37.980 |
| chr8 | 137135567 | rs78812419 | T | C | 39474 | 37.064 |

Supplementary table 14 Distribution of carriers, copies, and deceased patients of significantly associated variants grouped by individual populations, available at DOI: 10.5281/zenodo.24280139

